# Distinguishing Viruses Responsible for Influenza-Like Illness

**DOI:** 10.1101/2020.02.04.20020404

**Authors:** Julie A. Spencer, Deborah P. Shutt, Sarah K. Moser, Hannah Clegg, Helen J. Wearing, Harshini Mukundan, Carrie A. Manore

## Abstract

The many respiratory viruses that cause influenza-like illness (ILI) are reported and tracked as one entity, defined by the CDC as a group of symptoms that include a fever of 100 degrees Fahrenheit and a cough and/or a sore throat. In the United States alone, ILI impacts 9-49 million people every year. While tracking ILI as a single clinical syndrome is informative in many respects, the underlying viruses differ in their parameters and outbreak properties. Most existing models treat either a single respiratory virus or ILI as a whole. However, there is a need for models capable of comparing several individual ILI viruses. To address this need, here we present a flexible model and simulations of epidemics for influenza, RSV, rhinovirus, seasonal coronavirus, adenovirus, and SARS/MERS, parameterized by a systematic literature review and accompanied by a global sensitivity analysis. We find that for these biological causes of ILI, their parameter values, timing, prevalence, and proportional contributions differ substantially. These results demonstrate that distinguishing the viruses that cause influenza-like illness will be an important aspect of future work on ILI diagnostics, mitigation, modeling, and preparation for future unknown pandemics.

## 1. Introduction

Emerging infectious diseases are a major threat to global health security, as exemplified by the recent COVID-19 pandemic. The ease of transmissibility makes respiratory pathogens especially suited for epidemic spread [1]. Viral respiratory infections account for a large burden of annual morbidity and mortality worldwide [2] and are the cause more than 400,000 hospitalizations in children less than 18 years old [3] in the United States every year, demonstrating the perpetual scale of the challenge.

Most of these viral infections are categorized as Influenza-like Illnesses (ILI), which are defined as cases of possible influenza, or other illnesses resulting in a set of symptoms that are indistinguishable from those attributed to influenza viruses [4]. The CDC characterizes ILI as infections presenting with a fever of 100^º^F, and a cough and/or a sore throat [5], although common symptoms attributed to ILI include fever, chills, malaise, dry cough, loss of appetite, body aches, and nausea, combinations of which manifest depending on various pathogen-specific, environment specific, and host-determined factors [6].

The number of people impacted by ILI in the USA and beyond is significant every single year, notwithstanding the COVID pandemic. ILINet, which consists of outpatient healthcare providers in all 50 states, Puerto Rico, the District of Columbia and the U.S. Virgin Islands, reports over 60 million patient visits during the 2018-19 season [7, 8]. Indeed, 8% of the US population is considered to be infected with symptomatic influenza-like illness every year [9].

Defining ILI as a syndromic cluster rather than a specific disease or diseases is informative for keeping track of syndromic case counts, as well as for important analysis and forecasting [10]. However, the cluster of symptoms known as ILI is caused by many underlying pathogens [11, 12], most commonly, influenza viruses, common cold viruses, such as rhinovirus, adenovirus, human respiratory syncytial virus (RSV), parainfluenza virus (PIV), human metapneumovirus (hMPV) [13], and human coronaviruses (HCoV), a novel variant of which is responsible for the COVID-19 pandemic [14].

Despite the multifaceted biological etiology of ILI, diagnostic testing for specific viruses underlying ILI is relatively rare [5], and many of the diagnostic outcomes are based on syndromic evaluation at the point of care. There are no tailored discriminatory diagnostics for use at the point of care, to evaluate pathogens that impact 9-49 million people every year in the United States alone [5]. This creates a knowledge gap in which an emergent novel respiratory pathogen such as COVID-19 can go undetected [15]. An increased understanding of the biological dynamics of specific pathogens causing ILI is needed to prevent unnecessary suffering and death [16, 17, 18].

Although clinical studies have been conducted to assess the contribution of different viruses to ILI [19, 12, 20, 21, 22, 23], the reliance on syndromic diagnostics and the consequent impact on identification and of novel threats has not been assessed until recently [24]. Modeling studies that explore the mechanism of transmission and spread of ILI pathogens have also been conducted [25]. A recent study has shown that aggregrating the underlying ILI viruses separately rather than considering ILI as a single pathogen can improve ILI forecasts [24]. However, modeling studies using diagnostics measurements, and aimed at gaining insight into differing epidemic properties of the viruses underlying ILI, have been less abundant [26, 27].

To address the need for a flexible, abstract system that enables comparison of several ILI viruses in one paper, here we provide a deterministic model for five of the most common viral pathogens responsible for ILI. Our aim is to explore how pathogens with similar syndromes (and hence grouped together as ILI), can present with varied outbreak properties, thereby requiring varied interventions. We chose the pathogens on the basis of available literature, and the outcomes of a parallel clinical study conducted in Northern New Mexico Influenza (A and B), Respiratory Syncytial Virus (RSV), rhinovirus, Human Coronavirus (HCoV), and Adenovirus. We parameterized the model by conducting a systematic literature review, and aligned the associated sensitivity analysis with the gap analysis performed in our clinical studies.

This study presents a shift in perspective that contributes a practical foundation for advancement of diagnostics, interventions and improved pandemic preparedness of anticipated and novel ILI pathogens, and for developing modeling strategies that can support biosurveillance architectures and pandemic preparedness.

## 2. Methods

### 2.1. Model Structure

The equations governing this model of common upper respiratory virus dynamics are given by

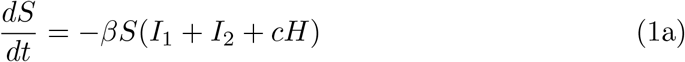

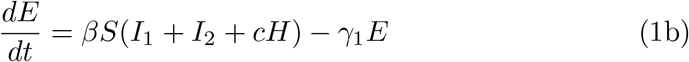

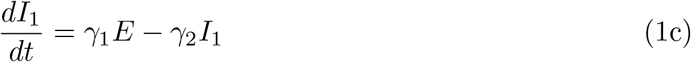

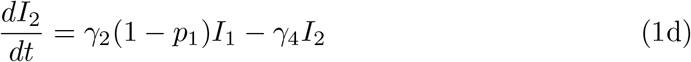

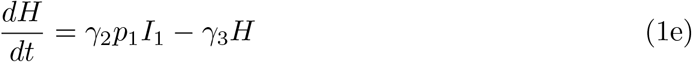

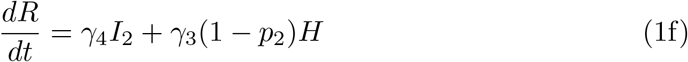

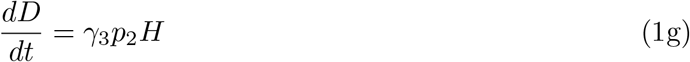

The total population is N = S + E + I_1_ + I_2_ + H + R + D.

To model five viruses that underly ILI during the course of one seasonal infection cycle, we developed a deterministic system of ordinary differential equations (Eq. 1). We then simulated daily infections of the five seasonal viruses after subtracting probable coinfections, and calculated the proportion of total daily infections contributed by each virus during the course of a hypothetical ILI season.

The model diagram (Fig. 1) illustrates the progression of ILI for 365 days in a human population of a hypothetical town containing 10,000 individuals. Five common seasonal ILI viruses, along with the historic outbreak coronaviruses SARS-CoV and MERS-Cov, are assumed to similar determininistic transmission structure.

**Figure 1:**
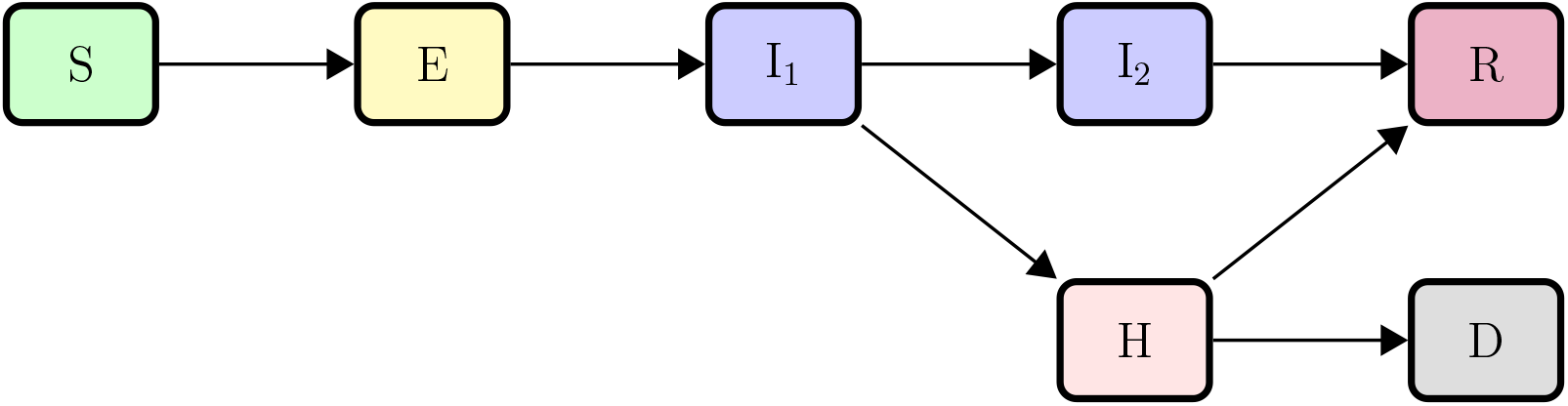
Transfer diagram for ILI virus transmission

The total population (N) consists of seven classes: susceptible (S), exposed but not infectious (E), first infectious class (I_1_), second (non-hospitalized) infectious class (I_2_), hospitalized (H), recovered (R), or deceased (D) (Table 1). Individuals are considered susceptible until they contact an infectious individual from (I_1_), (I_2_), or (H).

**Table 1:** Descriptions of state variables

| Variable | Description |
| --- | --- |
| $S$ | Number of susceptible individuals |
| $E$ | Number of exposed (not infectious) individuals |
| $I_1$ | Number of initially infectious individuals |
| $I_2$ | Number of infected, non-hospitalized individuals |
| $H$ | Number of hospitalized individuals |
| $R$ | Number of recovered individuals |
| $D$ | Number of deceased individuals |

We modeled the progression of disease as follows: Given contact with an infectious individual, transmission takes place with some probability. After transmission has occurred, susceptible people move to the exposed class (E), where they spend a number of days equal to the period between infection and the onset of infectiousness (the latent period). In accordance with accepted literature, we assume that the latent period equals the incubation period, which is the period of time between exposure to the virus and the onset of symptoms.

After the latent period, individuals move to the first infectious class, (I_1_). The duration of the first infectious period differs according to the underlying virus. Symptoms worsen for some proportion of individuals in the first infectious class, who then require hospitalization (H), where they remain infectious, with reduced transmission *c*. From the hospital, individuals either recover (R) or die (D). Individuals who remain sick, but do not require hospitalization for the duration of the second infectious period (I_2_), typically do not suffer from serious manifestations of the disease, and we assume that they recover entirely. We assume that hospitalized individuals have 25% less contact with susceptible individuals than do infectious people outside the hospital, which results in 25% less transmission during hospitalization. We further assume that all recovered individuals (R) gain full immunity to the virus causing the illness.

We assume that the total infected population (T = E + I_1_ + I_2_ + H) and the total infectious population (TI = I_1_ + I_2_ + H) are homogeneously mixed. We assume that in the duration of a single year, demographics (natural birth or death) are negligible, and they are not modeled. We assume that the viruses act independently, although coinfection is not ruled out. Each epidemic begins with a single infected individual, and we calculate the transmission rate *β* for each virus is by solving for *β* in the expression for *R*_0_ in Appendix B, using the mean *R*_0_ values for each virus from the literature.

### 2.2. Model Parameterization

To parameterize the model, we reviewed the literature for epidemiological measurements of incubation period, infectious period, hospitalization period, hospitalized proportion, case fatality proportion, and *R*_0_ (cf. Table 2) for influenza A and B, RSV, rhinoviruses, coronaviruses, and adenoviruses. We included results from experimental and observational studies, as well as from systematic reviews. We also included estimates of *R*_0_ from modeling studies, for even when symptomatically similar, many viral pathogens vary in their reproductive number. In the case of SARS, we included an estimate for the infectious period, since values were lacking in the literature [28]. We searched Google Scholar for each virus, using the name of the virus, a description of the parameter, and the type of study. For example, “influenza AND incubation period AND experimental” yielded a list of papers reporting the results of experimental studies to determine incubation period of influenza virus infections in humans. We then read the top 10 cited papers, examined the details of each study, and recorded the results (Table 3).

**Table 2:** Parameter descriptions and dimensions

| Parameter | Description | Dimension |
| --- | --- | --- |
| $\beta$ | basic transmission rate | $individuals^{-1} \times time^{-1}$ |
| $c$ | reduction of transmission in hospital | dimensionless |
| $\gamma_1$ | per capita rate of progress from exposed to infectious state | $time^{-1}$ |
| $\gamma_2$ | per capita rate of progress through initial infectious state | $time^{-1}$ |
| $\gamma_3$ | per capita rate of progress through hospitalized state | $time^{-1}$ |
| $\gamma_4$ | per capita rate of progress through non-hospitalized infectious state | $time^{-1}$ |
| $p_1$ | proportion of initially infectious population that becomes hospitalized | dimensionless |
| $p_2$ | proportion of hospitalized population that die | dimensionless |

**Table 3:** ILI parameters from literature. Coronavirus refer to the six pre-SARS-CoV-2 strains.

| Virus | Range<br>(Min to Max) | Mean | Standard<br>Deviation | References |
| --- | --- | --- | --- | --- |
| **Incubation Period (days)** |  |  |  |  |
| Influenza | 1-6.3 | 2.61 | 0.993 | [32, 33, 34, 35, 36, 37, 38, 39, 40] |
| RSV | 3-8 | 4.5 | 0.894 | [39, 32, 41, 42, 43, 40] |
| Rhinovirus | 0.42-5.5 | 2.36 | 1.10 | [39, 44, 45, 46, 47, 48, 49, 32, 43, 40] |
| Human coronavirus | 1.9-14.7 | 5.07 | 2.21 | [39, 50, 43, 40, 51, 52, 53, 54] |
| Adenovirus | 1-30 | 6.71 | 2.04 | [55, 39, 56, 57, 58, 40, 59] |
| **Infectious Period (days)** |  |  |  |  |
| Influenza | 1-9 | 4.58 | 2.56 | [33, 34, 37, 22, 60, 11] |
| RSV | 1-21 | 7.72 | 1.94 | [61, 62, 63, 11] |
| Rhinovirus | 7-16 | 9.40 | 1.70 | [11, 64, 65, 66, 67] |
| Human coronavirus | 7-35 | 15.20 | 10.30 | [11, 68, 69, 51, 54, 28] |
| Adenovirus | 7-17 | 8.20 | 2.89 | [55, 11, 70, 59] |
| **Hospitalization Period (days)** |  |  |  |  |
| Influenza | 3.5-11.3 | 6.36 | 3.27 | [22, 71, 72, 11, 73] |
| RSV | 2-17.5 | 5.24 | 2.32 | [74, 75, 76, 11, 77, 73] |
| Rhinovirus | 0.4-1.67 | 1.19 | 0.87 | [11, 78, 79] |
| Human coronavirus | 1.5-11 | 4.96 | 4.27 | [11, 69, 80, 81] |
| Adenovirus | 3.12-7 | 4.71 | 2.03 | [11, 77, 82] |
| **Hospitalization Proportion** |  |  |  |  |
| Influenza | 0.000035-0.062 | 0.0037 | 0.0075 | [83, 73, 84, 85, 11] |
| RSV | 0.00034-0.29 | 0.021 | 0.0215 | [86, 87, 83, 73, 88, 11, 62] |
| Rhinovirus | 0.0093-0.024 | 0.0121 | 0.0108 | [11, 89, 90] |
| Human coronavirus | 0.00224-0.52 | 0.188 | 0.241 | [11, 91, 92, 69] |
| Adenovirus | 0.014-0.95 | 0.43 | 0.39 | [12, 93, 11, 70] |
| **Case Fatality Proportion** |  |  |  |  |
| Influenza | 0.000106-0.0827 | 0.0312 | 0.0415 | [94, 95, 96, 97, 98] |
| RSV | 0.00031-0.165 | 0.0464 | 0.0627 | [99, 74, 95, 100, 88, 101] |
| Rhinovirus | 0-0.125 | 0.0451 | 0.0694 | [64, 102, 103] |
| Human coronavirus | 0-0.34 | 0.147 | 0.146 | [104, 105, 91, 92, 101, 103] |
| Adenovirus | 0.00075-0.166 | 0.103 | 0.0694 | [12, 106, 107, 108, 109, 70] |
| **R<sub>0</sub>** |  |  |  |  |
| Influenza | 1.06-3.4 | 1.68 | 0.871 | [110, 111, 112, 113, 114, 115] |
| RSV | 1.2-9.1 | 3.47 | 2.67 | [62, 116, 117, 118, 119, 120, 121] |
| Rhinovirus | 1.2-2.6 | 1.88 | 0.70 | [120, 121, 122] |
| Human coronavirus | 2.7-8 | 4.18 | 2.26 | [123, 105, 124, 125, 101, 126, 127, 128] |
| Adenovirus | 2.34 (one value) | 2.34 | NA | [26] |

We iterated this process for each pathogen, until either no additional studies could be found, or the information garnered had already been incorporated in our assessment. We consulted modeling and review studies only when experimental and observational studies were not available for a given pathogen. We obtained from two to nine values for each parameter, with the exceptions of *R*_0_ for Adenoviruses, and the hospitalization period for SARS and MERS, for which we found only one value each. We calculated the mean and standard deviation of each parameter (Table 3).

SARS-CoV-2, the virus that causes COVID-19, is a member of genus *Beta-coronavirus*, along with SARS-CoV and MERS-CoV [29]. Our parameter review includes values for strains 229E, NL63, OC43, HKU1, SARS-CoV, and MERS-CoV, the six commonly circulating strains of Coronaviruses that existed prior to the advent of COVID-19. We generated a separate Coronavirus parameter table, focused on comparing the parameter values of the seasonal strains to those of the more recent SARS-CoV and MERS-CoV (Table 4). We collected means when possible; and when means were not available, we recorded medians.

**Table 4:**
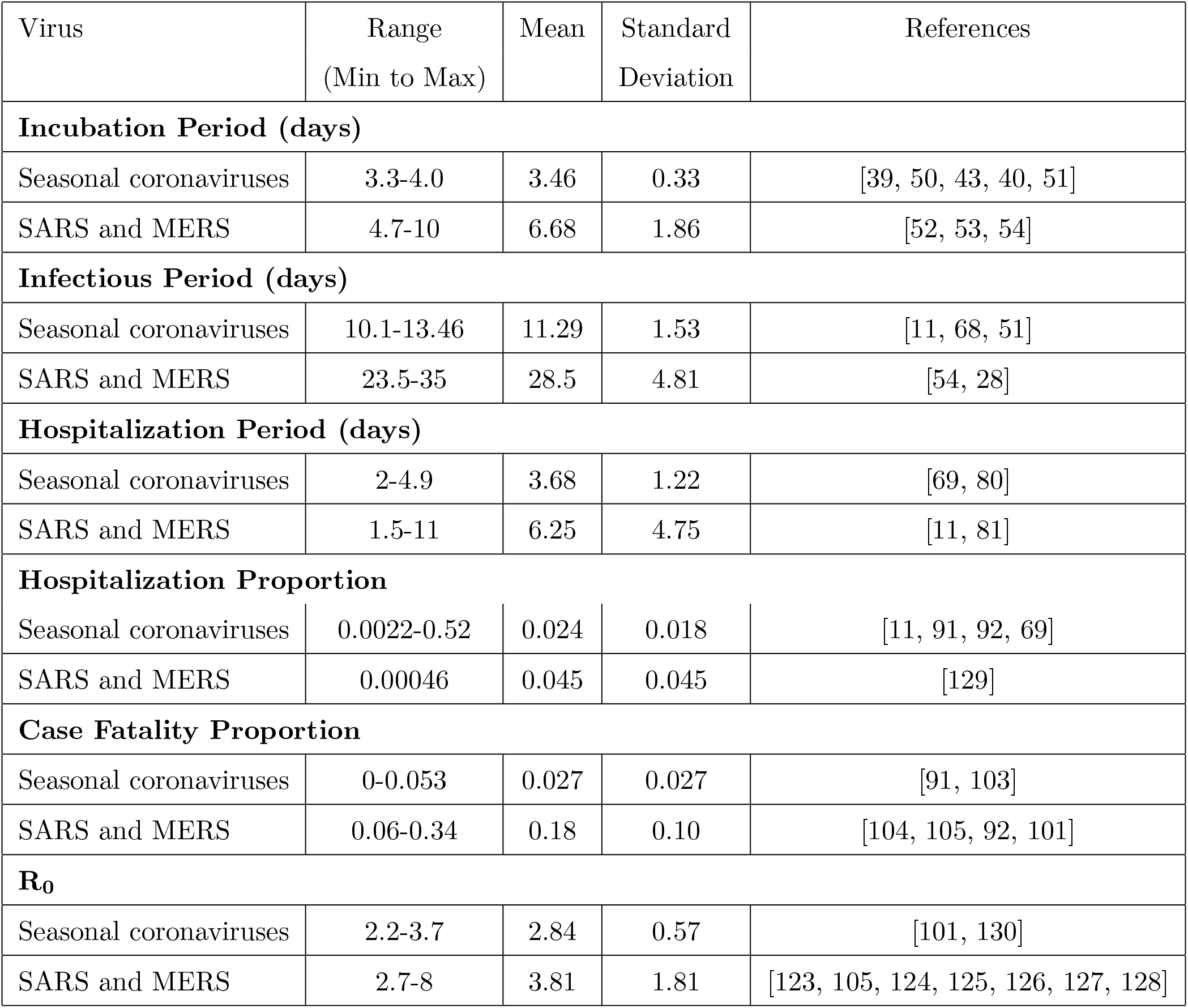
This table distinguishes between endemic seasonal coronaviruses (229E, NL63, OC43, and HKU1) and historic sporadic outbreak coronaviruses (SARS-CoV and MERS-CoV).

| Virus | Range<br>(Min to Max) | Mean | Standard<br>Deviation | References |
| --- | --- | --- | --- | --- |
| **Incubation Period (days)** |  |  |  |  |
| Seasonal coronaviruses | 3.3-4.0 | 3.46 | 0.33 | [39, 50, 43, 40, 51] |
| SARS and MERS | 4.7-10 | 6.68 | 1.86 | [52, 53, 54] |
| **Infectious Period (days)** |  |  |  |  |
| Seasonal coronaviruses | 10.1-13.46 | 11.29 | 1.53 | [11, 68, 51] |
| SARS and MERS | 23.5-35 | 28.5 | 4.81 | [54, 28] |
| **Hospitalization Period (days)** |  |  |  |  |
| Seasonal coronaviruses | 2-4.9 | 3.68 | 1.22 | [69, 80] |
| SARS and MERS | 1.5-11 | 6.25 | 4.75 | [11, 81] |
| **Hospitalization Proportion** |  |  |  |  |
| Seasonal coronaviruses | 0.0022-0.52 | 0.024 | 0.018 | [11, 91, 92, 69] |
| SARS and MERS | 0.00046 | 0.045 | 0.045 | [129] |
| **Case Fatality Proportion** |  |  |  |  |
| Seasonal coronaviruses | 0-0.053 | 0.027 | 0.027 | [91, 103] |
| SARS and MERS | 0.06-0.34 | 0.18 | 0.10 | [104, 105, 92, 101] |
| **R<sub>0</sub>** |  |  |  |  |
| Seasonal coronaviruses | 2.2-3.7 | 2.84 | 0.57 | [101, 130] |
| SARS and MERS | 2.7-8 | 3.81 | 1.81 | [123, 105, 124, 125, 126, 127, 128] |

### 2.3. Global Sensitivity Analysis

To prioritize the impact of parameters on model outputs for this nonlinear system, we carried out a global sensitivity analysis. We bounded the parameter space with the minimum and maximum parameters from the literature (Tables 3, 4, Appendix C.4). We simulated epidemics of five common upper respiratory viruses implicated in ILI: influenza, respiratory syncytial virus (RSV), Rhinovirus, seasonal human coronavirus (HCoV), and adenovirus, alongside outbreak strains of SARS-CoV and MERS-CoV, grouped together.

We assessed the impact of five model input variables of *β* (basic transmission rate), *γ*_1_ (1/incubation period), *γ*_2_ (1/onset to hospitalization), *γ*_3_ (1/hospitalized period), and *γ*_4_ (1/non-hospitalized period) on three response variables of total number of infections, magnitude of epidemic peak (peak height), and time to epidemic peak. We used Latin Hypercube Sampling (LHS) [30] to generate 10,000 sets of values from the total parameter space for each of the five parameters, for each of the viruses. We considered the parameter ranges for seasonal coronaviruses (HCoV) separately from SARS-CoV/MERS-CoV. We calculated *β* ranges by solving Equation B.4 for *β* and substituting the minimum and maximum values from the literature (Table D.5). In the case of adenovirus, we found only one *R*_0_ value in the literature, and assumed minimum and maximum values of −20% and +20% from this single value. We solved the ODE system numerically for these sample input values using the default integration routine “ode” in R package deSolve, then constructed a dataframe of the marginal relations between the individual parameters and outputs. We generated sensitivity plots in R using Smoothed Conditional Means (“statsmooth”) and trend lines using weighted least squares method Local Polynomial Regression Fitting in R (“loess”) [31]. We assumed a uniform distribution on the parameters.

## 3. Results

### 3.1. Model Simulation Results

We find that when we numerically simulate seasonal epidemics for five common ILI viruses, setting all of their starting times at October 1st, the varied ranges of historic parameter values for each virus result in varied timing, prevalence, and contributions of these underlying biological causes of ILI (Figure 2). RSV peaks in December, followed by rhinovirus. Seasonal coronavirus peaks in January, adenovirus peaks in February, and influenza peaks in March. RSV has the greatest total cumulative infections and the highest peak (greatest maximum daily number of infections), while influenza has the least total cumulative infections and the lowest peak. The numerical simulation of SARS and MERS, not illustrated here as an epidemic curve because not considered seasonal, has the greatest overall number of cumulative infections.

**Figure 2:**
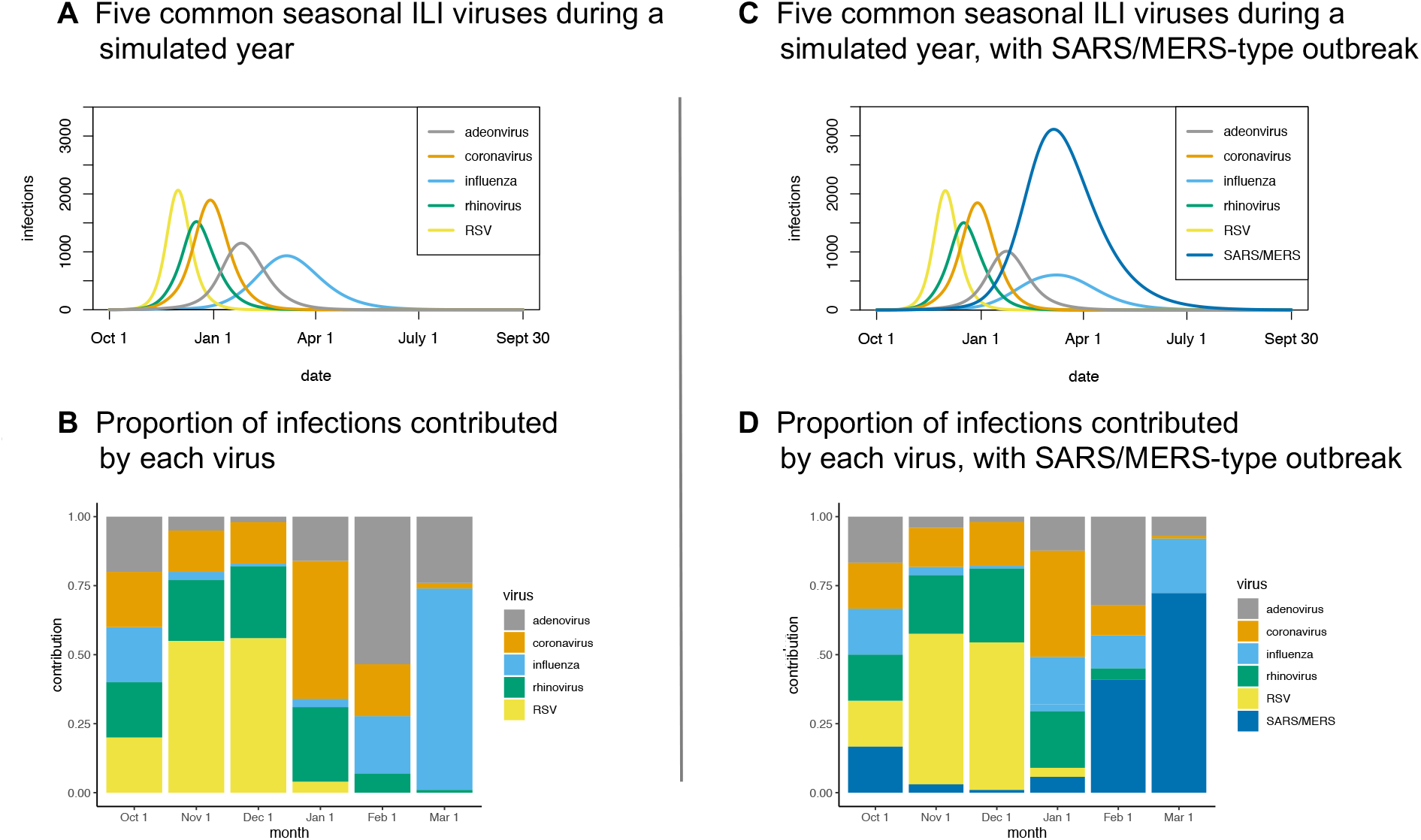
Seasonal and outbreak simulations. Panel 2A displays a numerical simulation of daily infections for five seasonal ILI viruses for one hypothetical year. The x-axis shows time in days; the y-axis shows number of infections. Each virus begins by infecting one individual, with 9,999 susceptible individuals. We assume no background immunity, vaccination, or mitigations; each virus acts independently; probable coinfections are subtracted. Inputted parameter values are the means from the literature (Table 3). Values for coronavirus are the means for the pre-SARS-CoV-2, endemic, seasonal coronaviruses OC43, 229E, HKU1, and NL63, considered as a group. Panel 2B displays six snapshots of the proportion contributed by each virus on the first day of each month of the hypothetical “flu” season. The x-axis shows the first day of each month; the y-axis shows the proportional contributions. Panels 2C and 2D are the analogous plots, with the inclusion of the outbreak coronaviruses SARS-CoV and MERS-CoV, considered as a group.

We find that on the first day of any given month during the simulated ILI season, the composition of ILI attributable to the individual viruses varies considerably (see the stacked bar chart in (Figure 2)). On October 1st, each represents 20% of the total five infections, reflecting the initial condition that each epidemic begins with one infection. On November 1st and December 1st respectively, RSV constitutes 55% and 56% of total ILI infections. On January 1st, seasonal coronavirus contributes 50% of the total; on February 1st, adenovirus contributes 54%; on March 1st, influenza contributes 73%.

### 3.2. Model Parameterization Results

We find 104 studies that contained relevant parameter values (Table 3). According to our literature review, adenovirus exhibits the longest mean incubation period, seasonal coronavirus has the longest total mean infection period, and RSV has the highest mean *R*_0_. Notably, out of the viruses studied, RSV incubation period values have the least standard deviation, while adenovirus values have the greatest standard deviation. Rhinovirus infectious period values have the least standard deviation, while SARS/MERS has the greatest. And finally, rhinovirus *R*_0_ values have the least variation, while RSV *R*_0_ values have the greatest variation.

We find that the seasonal and historical outbreak coronaviruses differ considerably in their defining epidemiological parameters. The incubation and hospitalization periods of SARS and MERS are almost double that of their seasonal counterparts, while their infectious period is more than double. The mean *R*_0_ for the betacoronaviruses SARS and MERS is 3.81, while that of the seasonal coronavirus strains is 2.84 (Table 4).

Of the 104 studies, 10 provide values for the percentage of each of the five viruses in question identified among ILI patients. In these 10 studies, on average, at least one virus is identified in 62% of individuals with ILI symptoms. Out of these 62% (mean values), influenza is identified in a 21.3% of samples, RSV in 13.5%, rhinovirus in 22.6%, human coronavirus in 8.8%, and adenovirus in 8.1%. The presence or absence of co-infection is not taken into account in the percentages reported by these studies.

### 3.3. Global Sensitivity Results

The global sensitivity analysis for our nonlinear system reveals several differences between the six viruses under consideration: influenza, RSV, rhinovirus, seasonal coronavirus, adenovirus, and SARS/MERS.

The impact of the basic transmission rate *β* on total cumulative infections is relatively tightly constrained for all six viruses (Appendix E). The trend line for influenza is slightly sigmoid, while the trend lines appear exponential for the other five, approaching 100% of the population asymptotically near their mean *β* values. For influenza and RSV, the Latin Hypercube Samples (LHS) for all five input variables have bimodal distributions for cumulative cases and time to peak. That is, for influenza and RSV, but not for the other four viruses, there are trivial numerical solutions for the system in which epidemics never take off, as well as nontrivial solutions, where they do take off. This is reflected in the disease-free equilibrium (Appendix A) and in the histograms (Appendix E).

The impact of *β* on peak height for the 10,000 LHS samples is not as constrained overall as for cumulative cases, although the variance is narrower than that of the other four input variables. The LHS sample distributions for output variable time to peak are bimodal for influenza and RSV, while they are unimodal for the other four viruses. Distributions for seasonal coronaviruses and historic outbreak coronaviruses (SARS/MERS) are unimodal (Appendix E).

An important difference between the seasonal and outbreak coronaviruses may be seen in Figure 3. Note that the y-axis for the outbreak coronaviruses represents approximately double the time period of the y-axis for seasonal coronaviruses. In both cases, the relationship of the transmission rate to the number of days to the epidemic peak is logarithmic and well constrained. In both cases, as the transmission rate increases, the time to peak shortens; conversely, as the transmission rate decreases, the time to peak is delayed. The other four input variables, representing speed of progression through the stages of disease, have unremarkable impacts on the output variables.

**Figure 3:**
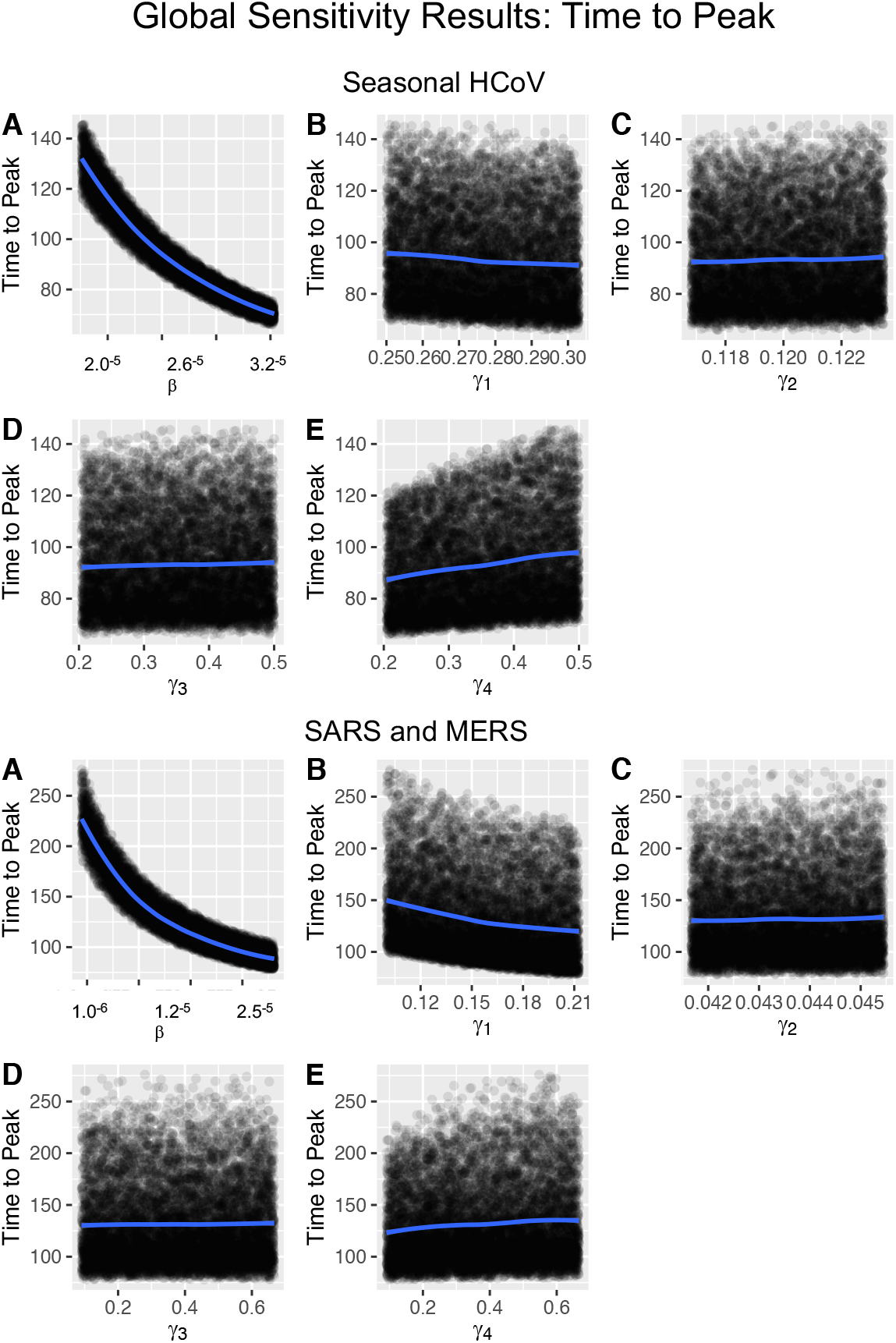
Global sensitivity analysis for seasonal coronaviruses (top) and historic outbreak coronaviruses SARS and MERS (bottom), showing the impact of five input variables on model output variable of time to epidemic peak. Note the different scale of the y axes on the *β* plots. Complete sensitivity results may be found in Appendix E.

## 4. Discussion

Although the five viruses considered here present clinically with similar symptoms, their parameters and epidemic characteristics differ, illustrating the vast potential not only for misdiagnosis and uninformed mitigation strategies, but for missing early signals of future novel emerging diseases. These results support those in Pei and Shaman’s recent paper [24] that demonstate differing outbreak properties of individual viruses that contribute to ILI.

From our sensitivity analysis, it is apparent that virus transmission rates, and therefore also effective reproduction numbers, have much greater impact on the output variables than do the other four input variables, more biologically intrinsic to each virus, that represent speed of the phases of disease progression. From a mitigation perspective, the transmission rate is the variable we can impact with public health policies. For example, by implementing mask-wearing and physical distancing, we can reduce and delay epidemic peak(s) of outbreak respiratory viruses, reducing cases and deaths as well as the burden on the healthcare system.

Many of the studies that generated parameter values evaluated populations treated at clinics or admitted at hospitals. However, a significant proportion of illness and death may occur outside of hospitals and clinics (see Cohen et al. 2017). Our formula for *R*_0_ is based on the traditional assumption of a naïve population; however, our parameter review reports many values from clinical studies conducted on populations that have been exposed to the seasonal ILI viruses in circulation, and therefore have some level of background immunity. Thus, these values likely reflect the effective reproductive number (*R*_*e*_) rather than *R*_0_, with the exception of SARS-CoV and MERS-CoV, which were novel zoonoses when they appeared. Further, the studies reviewed herein have taken place over many decades, during which viral evolution may be presumed to have occurred.

In our simulations, we treat the viruses contributing to the clinical syndrome of influenza-like illness as though they act independently, although a recent theoretical study has shown that even noninteracting pathogens are not necessarily mathematically independent [131]. A recent clinical study has shown rhinovirus can function to block subsequent infection by influenza [132]. Further studies are needed to assess the potential for both within-host and between-host viral interactions.

A limitation of our simulation results is that we have based them on mean historic parameter values, which does not take into account seasonal impacts on those parameters. Further, we have uniformly introduced the first infected individual for all viruses on October 1st of our hypothetical ILI season; however, in real world phenomena, these initial infections may happen at different times in different populations, and could happen in varied orders in different years.

The flexible deterministic model, numerical simulations, and sensitivity analysis included herein set the stage for future studies to investigate potential interactions of individual viruses that contribute to ILI. The parameterization study provides a meta-analysis of clinical studies during the past century that have provided the basic epidemiological parameters for modeling five of the common viruses contributing to the clinical syndrome of influenza-like illness. Along with previous work, the results presented herein indicate that in order to improve diagnosis, mitigation, and modeling of respiratory viruses, as well as to be prepared for the next pandemic, individual viruses contributing to influenza-like illness should be considered separately.

### CREdiT authorship contribution statement

**Julie A. Spencer:** Conceptualization, Methodology, Formal analysis, Visualization, Writing - Original Draft, Writing - Review & Editing. **Deborah P. Shutt:** Conceptualization, Methodology. **Sarah K. Moser:** Investigation, Visualization. **Hannah Clegg:** Conceptualization, Investigation. **Helen J. Wearing:** Conceptualization, Methodology, Formal analysis, Supervision, Writing - Review & Editing. **Harshini Mukundan:** Conceptualization, Project administration, Funding acquisition, Supervision, Writing - Review & Editing. **Carrie A. Manore:** Conceptualization, Methodology, Funding acquisition, Supervision, Writing - Review & Editing.

## Data Availability

The spreadsheets that contain the details of our parameter review are included in the paper, after the reference section.

## Appendix A. Disease-free Equilibrium

In the disease-free state, all infected classes are zero, that is, *E* = *I*_1_ = *I*_2_ = *H* = 0. Substituting and setting the derivatives equal to zero, it is evident that in the disease-free state, the other state variables R and D will continue to contain zero individuals, and that the Susceptible class S will remain equal to the total population N.

If we set any one of E, I_1_, I_2_, or H to zero, the other three state variables representing infected classes must also be zero. In this case, N=S=10000. Thus, where *x* = (*S, E, I*_1_, *I*_2_, *H, R, D*) denotes solutions of the system, *x*_*dfe*_ = (10000, 0, 0, 0, 0, 0, 0) represents the disease-free equilibrium for the system.

## Appendix B. Derivation of Basic Reproductive Number

The basic reproductive number (*R*_0_) is defined as the average number of secondary infections produced when one infected individual is introduced into a fully susceptible population. Four compartments, latently infected individuals (E), symptomatic and infected individuals (I_1_), symptomatic and infected and non-hospitalized individuals (I_2_), and hospitalized individuals (H), together characterize the total infected population for the ILI virus system. To calculate *R*_0_ for this system, we derive the next generation matrix [133].

Method:

1. Derive the matrix for the transmission term describing everyone entering (E): the “F” matrix;
2. Derive the matrix for the transition terms describing everyone transitioning between infected classes (*E, I*_1_, *I*_2_, *H*): the “V” matrix;
3. Next Generation Matrix (NGM) = (*F*)(*V* ^−1^);
4. The largest dominant eigenvalue or spectral radius of the NGM = *R*_0_ for the system.

The transmission term for the system is *βS*(*I*_1_ + *I*_2_ + *cH*)

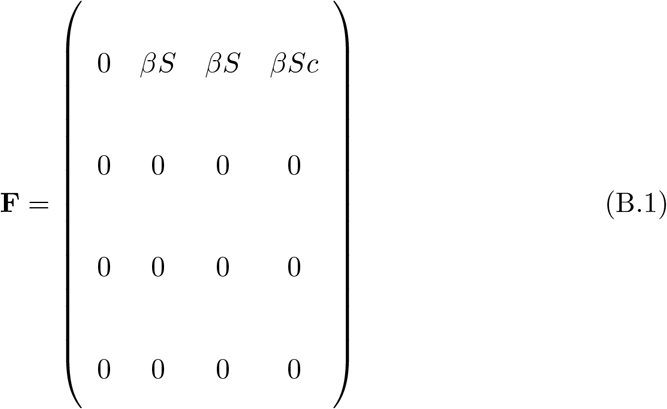

The transition terms for the system are (−*γ*_1_*E*), (*γ*_1_*E* − *γ*_2_*I*_1_), (*γ*_2_(1 − *p*_1_)*I*_1_ − *γ*_4_*I*_2_), (*γ*_2_*p*_1_*I*_1_ − *γ*_3_*H*).

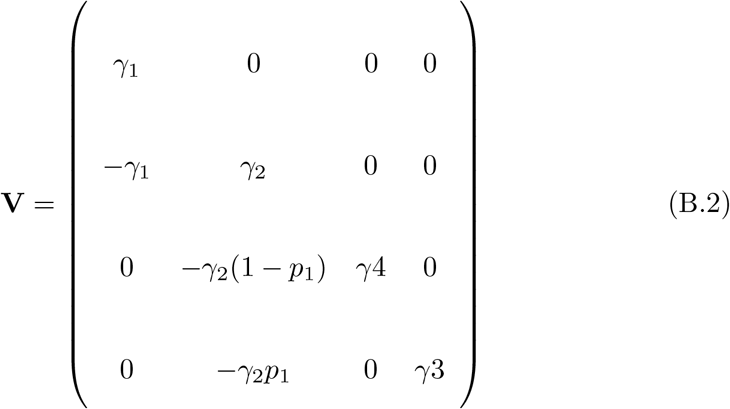

The next generation matrix is thus

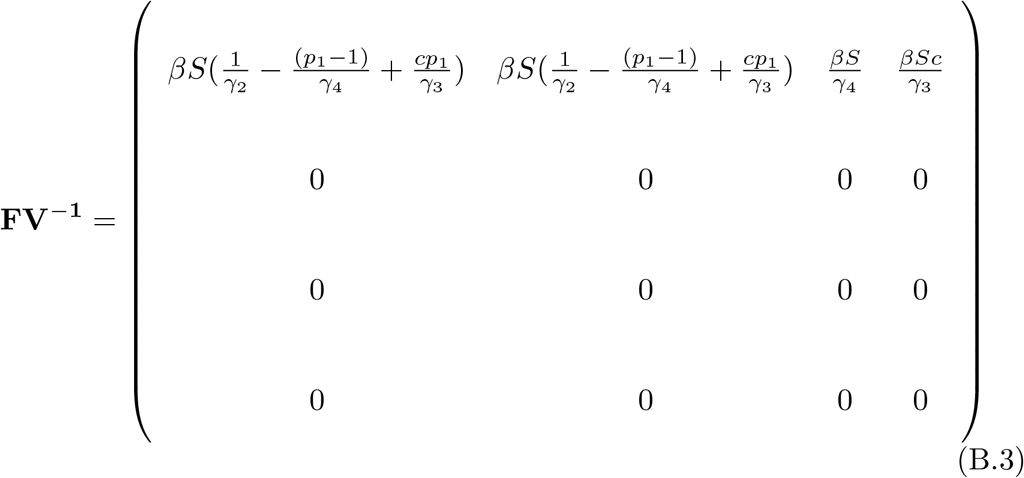

The spectral radius, or the largest positive eigenvalue of the next generation matrix, is the basic reproductive number of the system at the disease-free equilibrium.

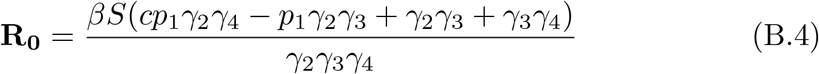

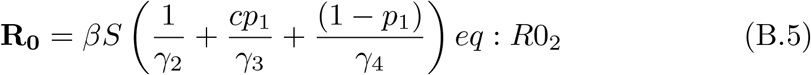

This expression for the basic reproductive number (*R*_0_) depends on the parameters *β, c, p*_1_, *γ*_2_, *γ*_3_ and *γ*_4_, and on the initial conditions of the state variables. Equation B.5 shows that *R*_0_ for this system is a combination of the transmission that takes place in the pre-symptomatic (*I*_1_), symptomatic, (*I*_2_), and hospitalized (H) compartments. This is the per-day transmission rate (*β*) multiplied by the time spent in each of these compartments.

## Appendix C. Parameter Ranges

**Figure C.4:**
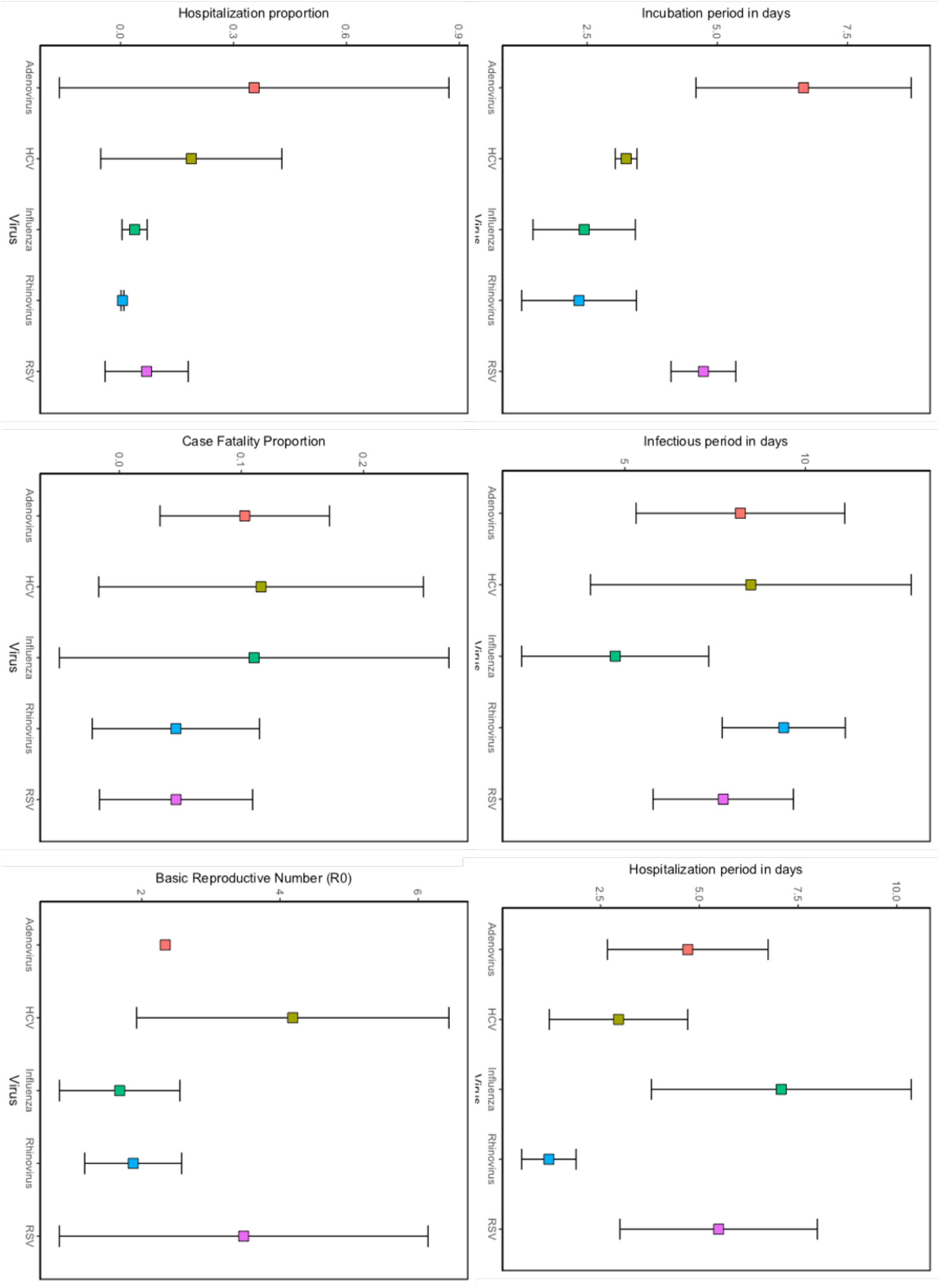
Parameter ranges for five common ILI viruses from literature review

## Appendix D. Transmission Rates

**Table D.5:** This table reports *β* (basic transmission rate) values calculated from solving Equation (6) for *β*, and substituting in the parameter values from the literature review (Tables 3 and 4).

| Virus | Min | Max | Mean |
| --- | --- | --- | --- |
| Influenza | $9.69^{-6}$ | $3.12^{-5}$ | $1.54^{-5}$ |
| RSV | $1.55^{-5}$ | $1.18^{-4}$ | $4.49^{-5}$ |
| Rhinovirus | $1.81^{-5}$ | $3.92^{-5}$ | $2.83^{-5}$ |
| Seasonal HCoV | $1.95^{-5}$ | $3.28^{-5}$ | $2.52^{-5}$ |
| Adenovirus | $2.28^{-5}$ | $3.42^{-5}$ | $2.85^{-5}$ |
| SARS/MERS | $9.47^{-6}$ | $2.81^{-5}$ | $1.34^{-5}$ |

## Appendix F. References

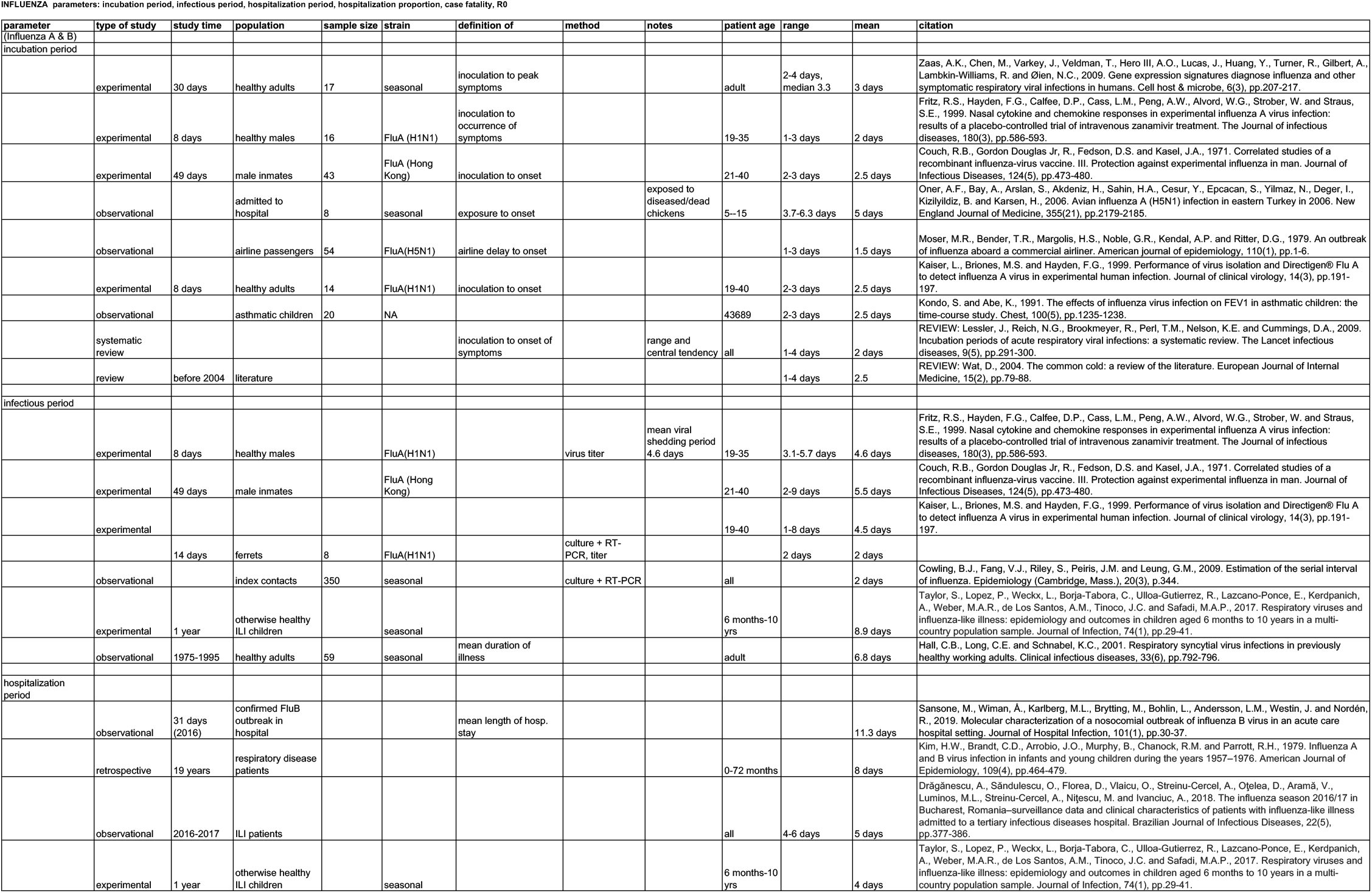

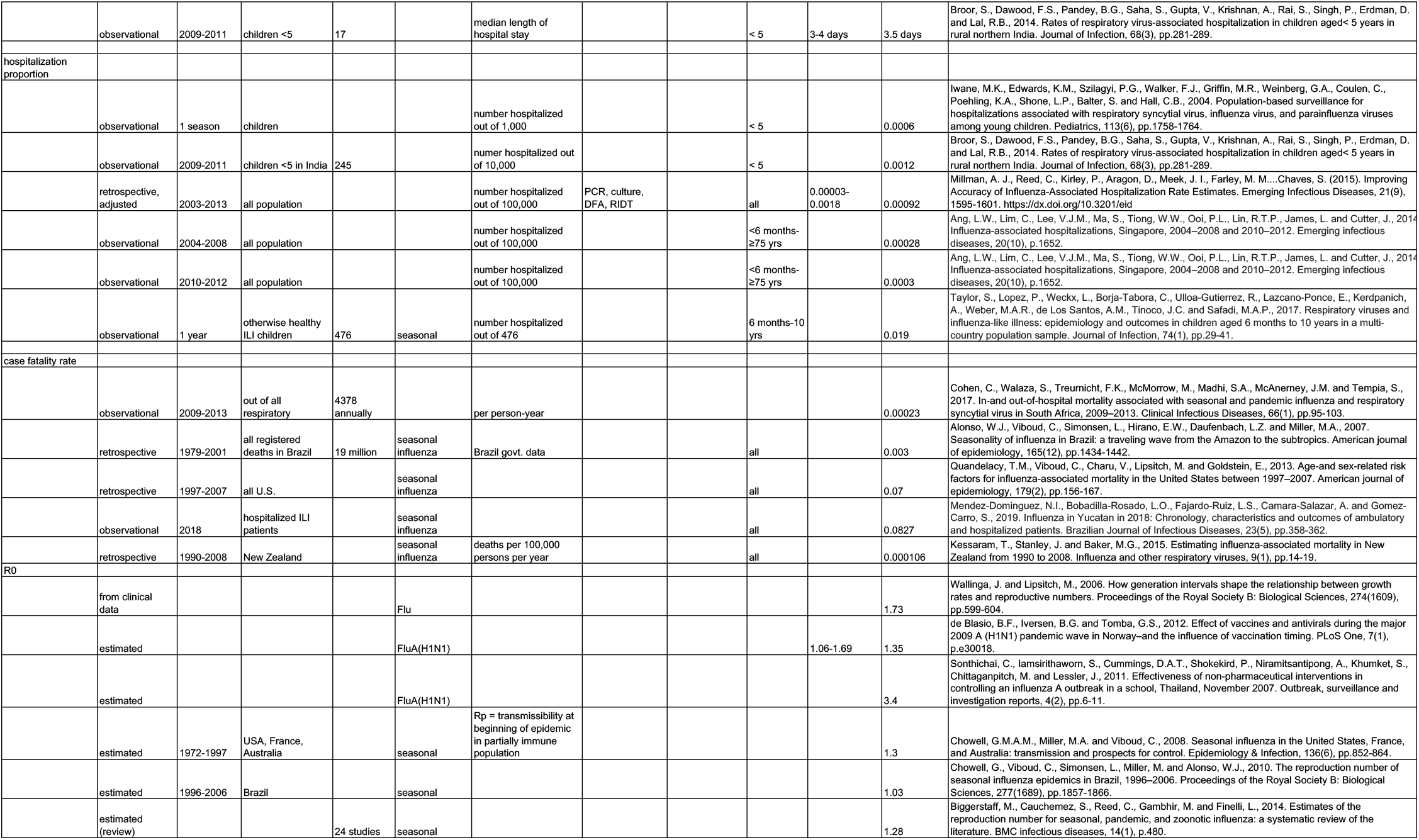

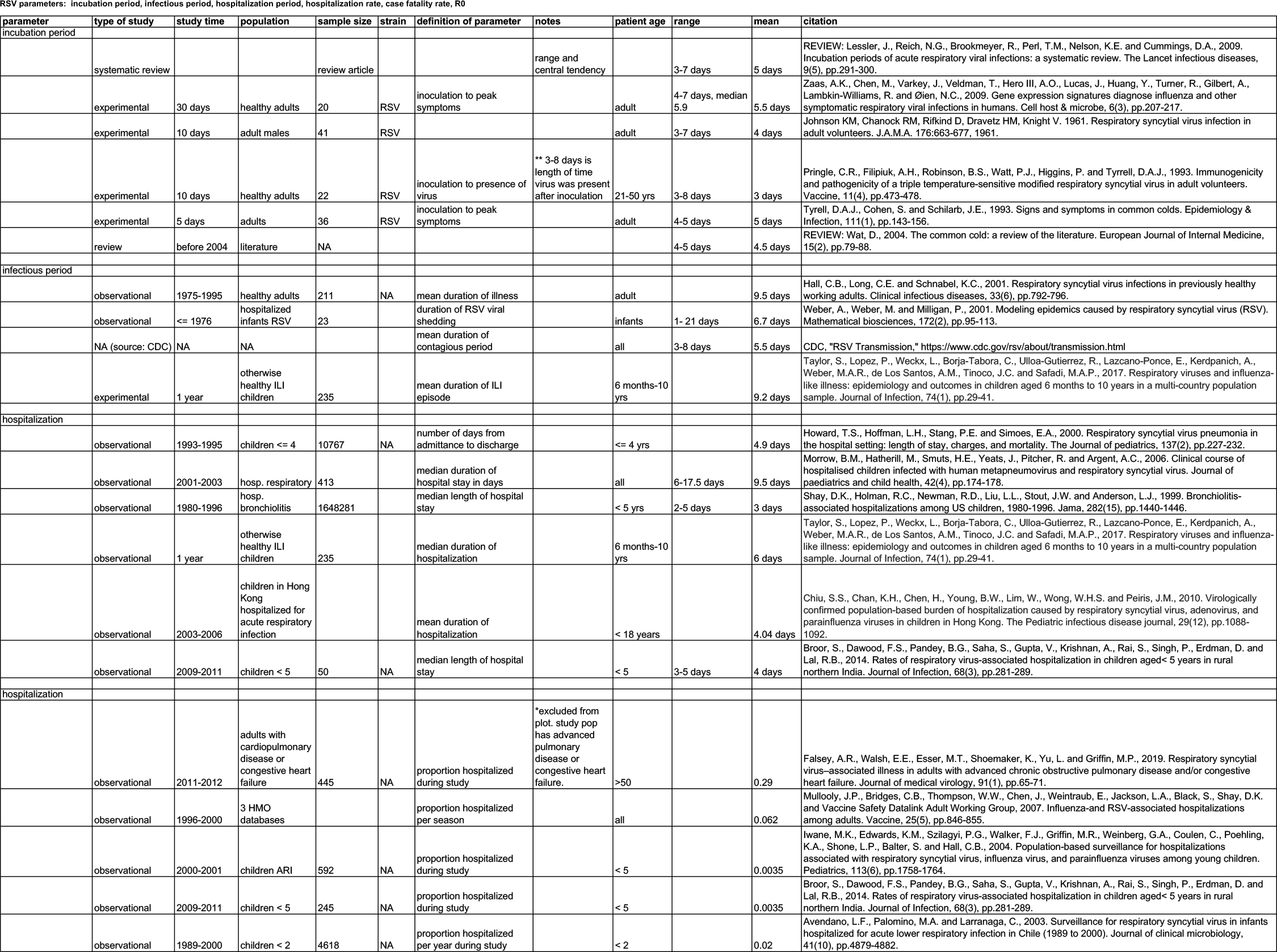

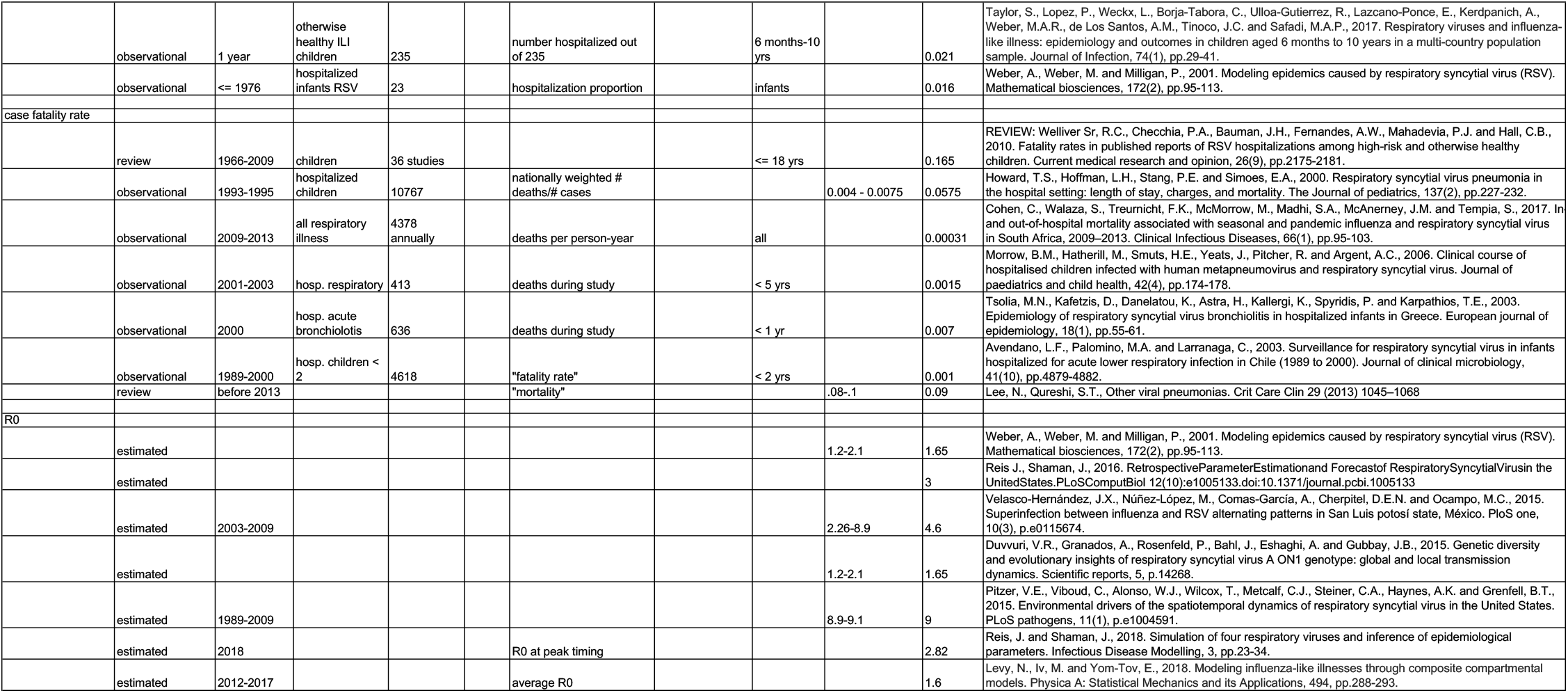

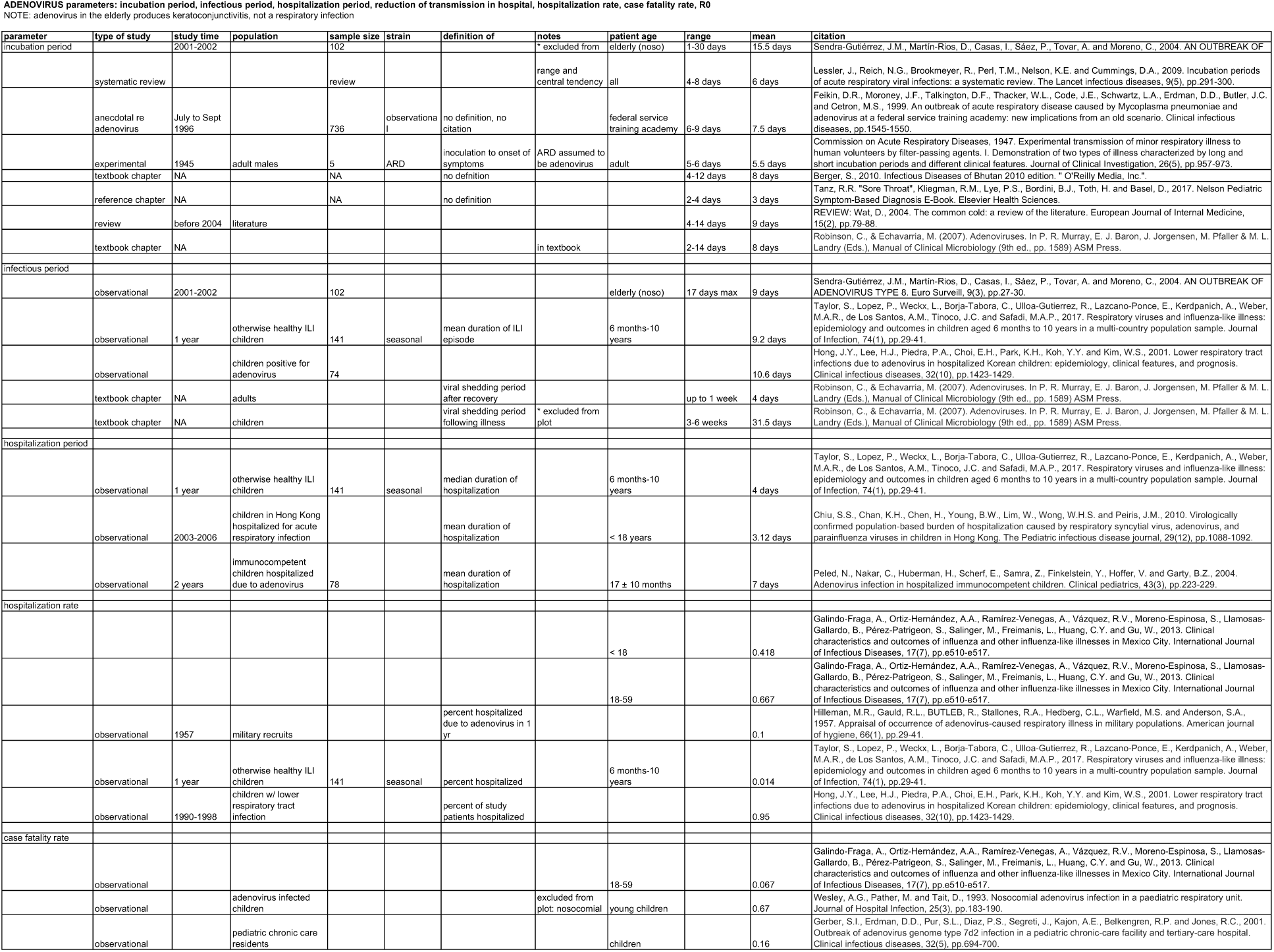

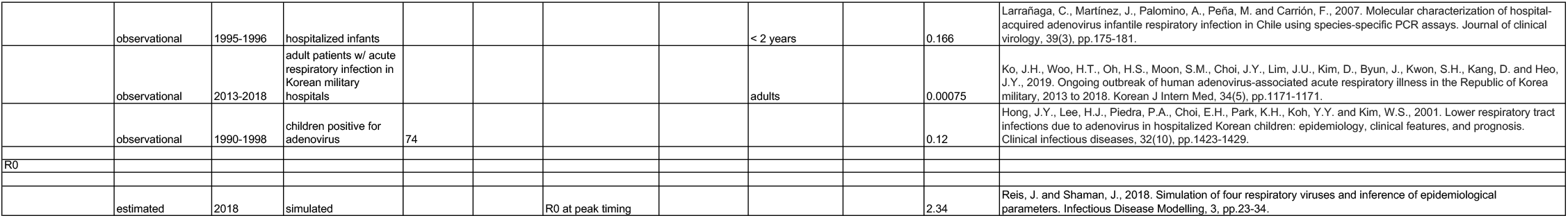

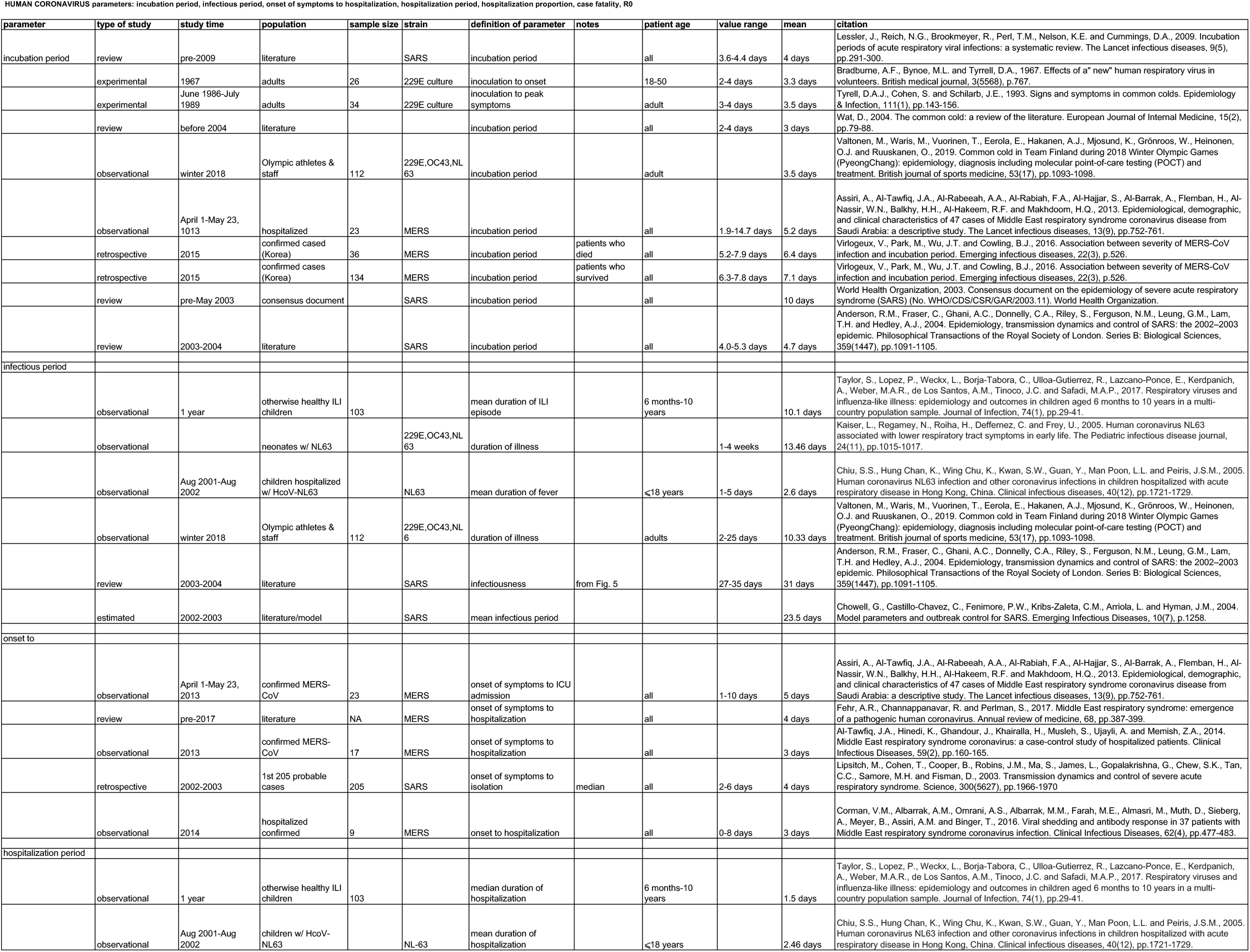

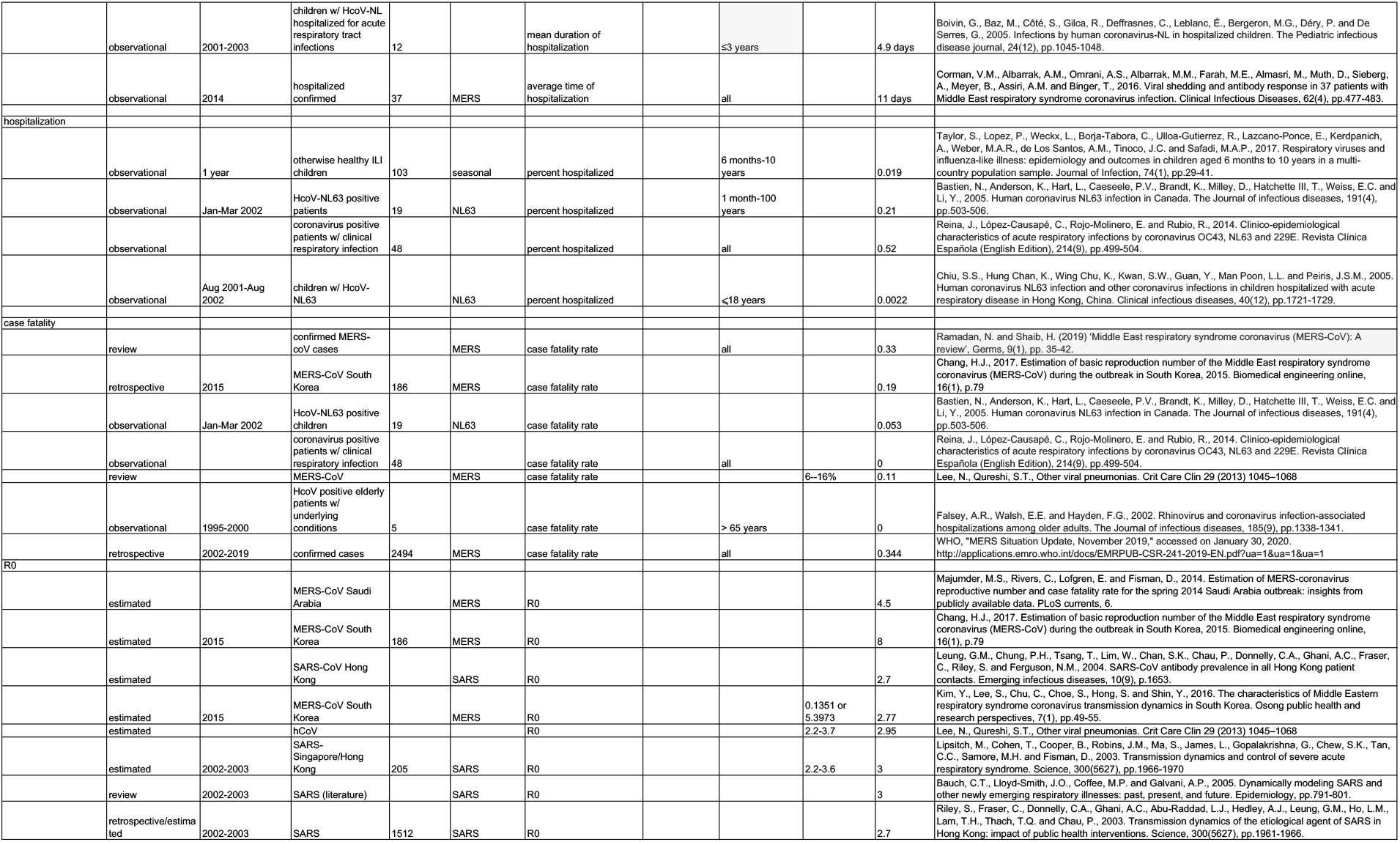

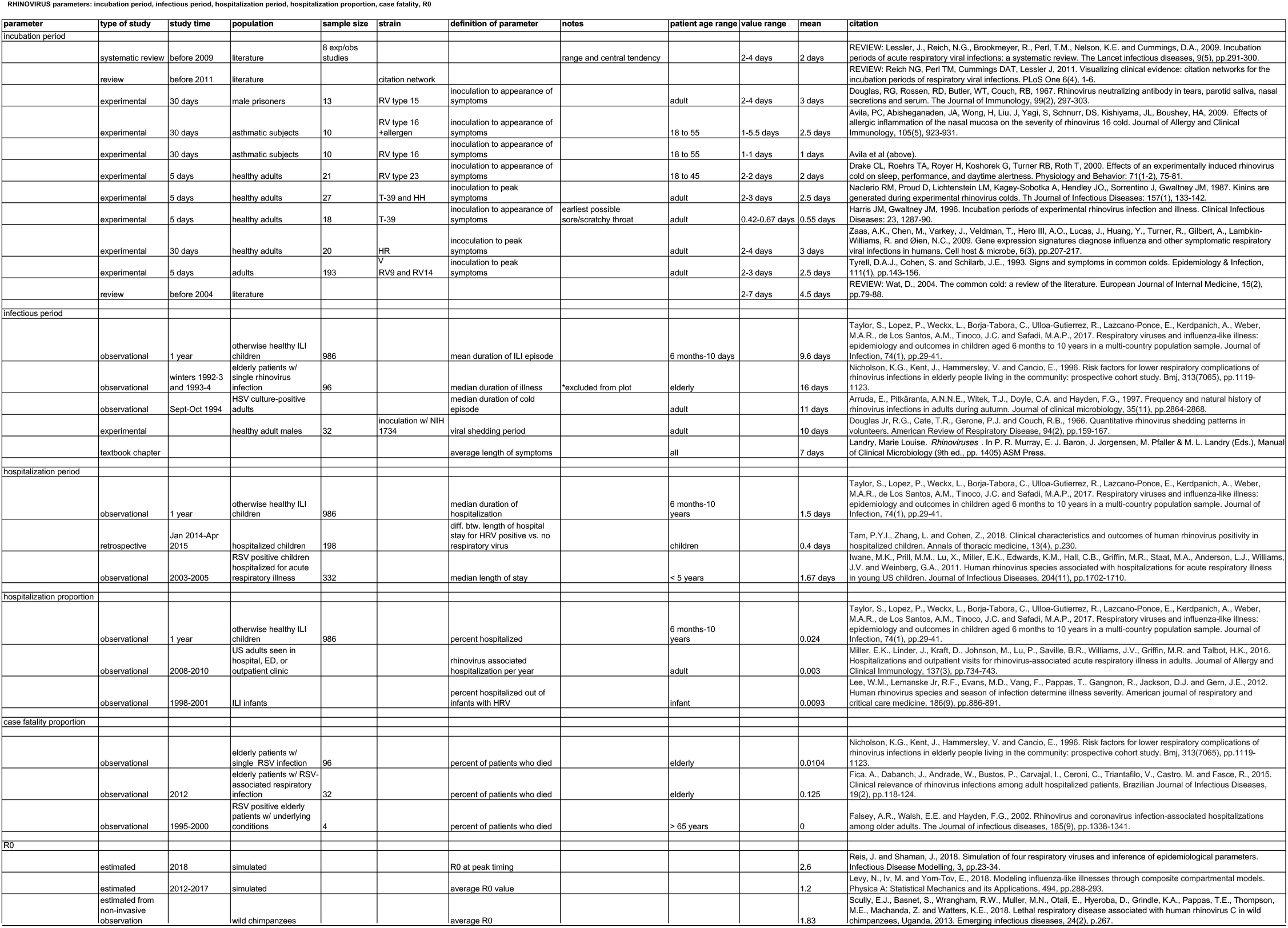

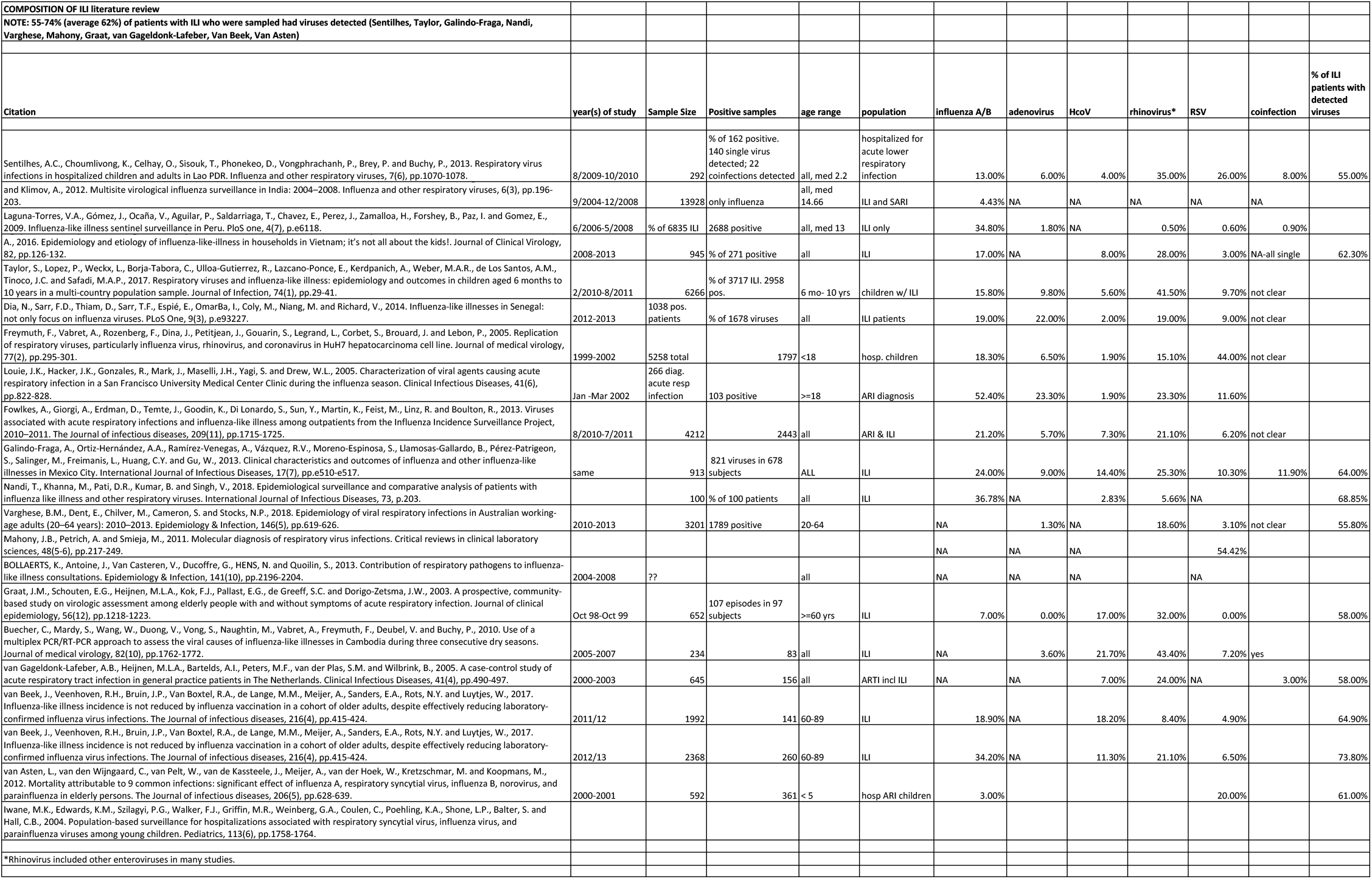

