## Appendix E for "Distinguishing Viruses Responsible for Influenza-Like Illness"

### Appendix E. Global Sensitivity Plots

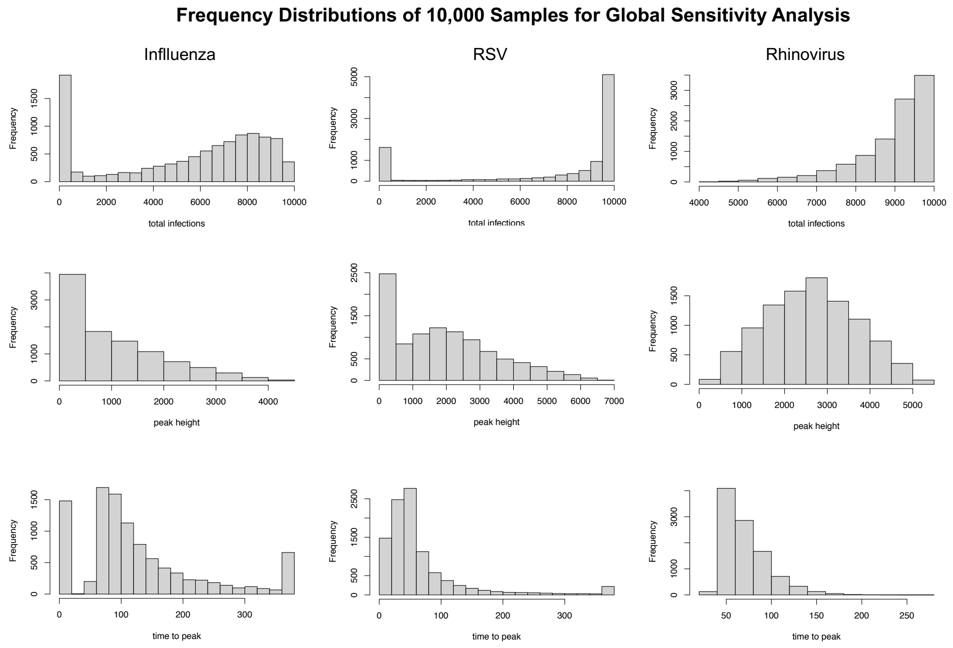


Figure E.5: Frequency Distributions of response variables total infections (top row), epidemic peak height (middle row), and number of days to epidemic peak (bottom row), from global sensitivity analysis for influenza, RSV, and rhinovirus.

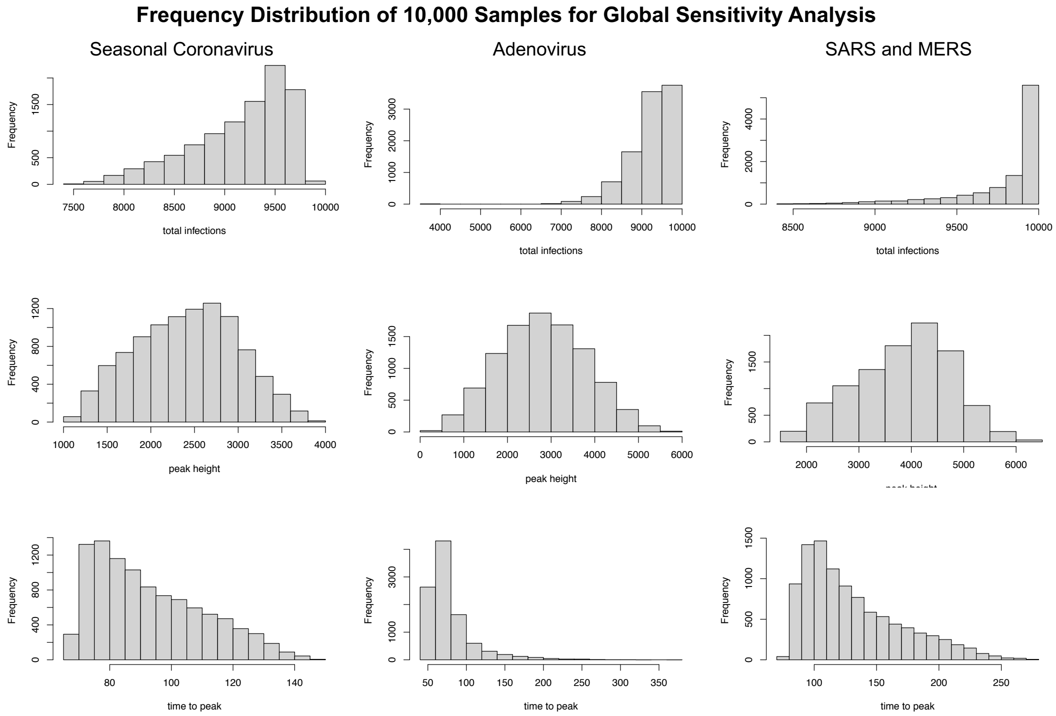


Figure E.6: Frequency Distributions of response variables total infections (top row), epidemic peak height (middle row), and number of days to epidemic peak (bottom row), from global sensitivity analysis for seasonal coronaviruses, adenovirus, and SARS/MERS.

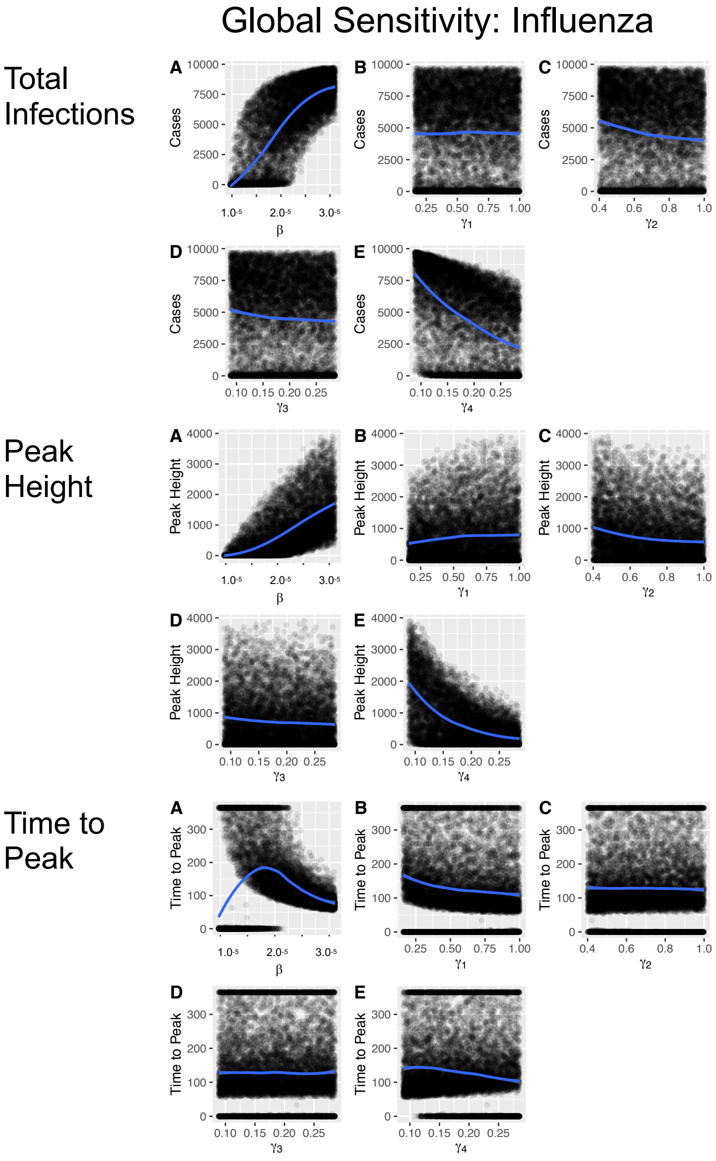


Figure E.7: Global sensitivity analysis for influenza. Subplots show the distribution of 10,000 Latin Hypercube Samples for input variables $\beta, \gamma_1, \gamma_2, \gamma_3$, and $\gamma_4$ respectively. Local Polynomial Regression trend lines show their relationship to response variables total infections at the top group, peak height in the middle group, and days to peak at the bottom group.

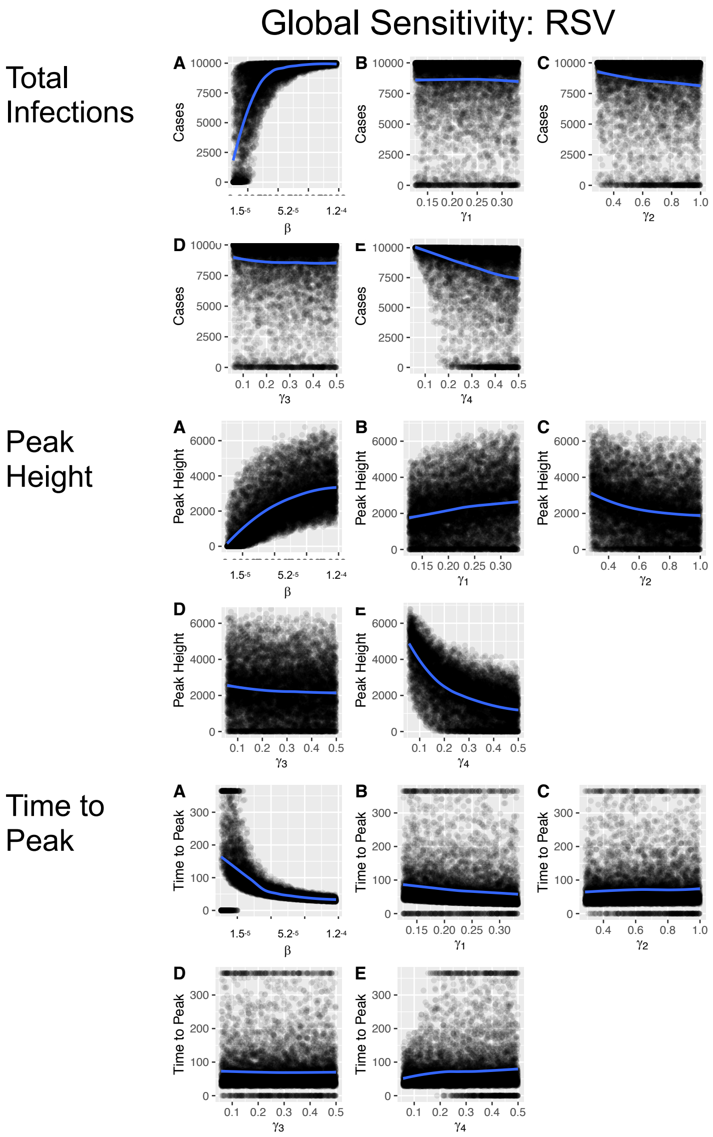


Figure E.8: Global sensitivity analysis for RSV. Subplots A, B, C, D, and E show the distribution of 10,000 Latin Hypercube Samples for input variables $\beta, \gamma_1, \gamma_2, \gamma_3$, and $\gamma_4$ respectively. Local Polynomial Regression trend lines show their relationship to response variables total infections at the top, peak height in the middle, and time to peak at the bottom.

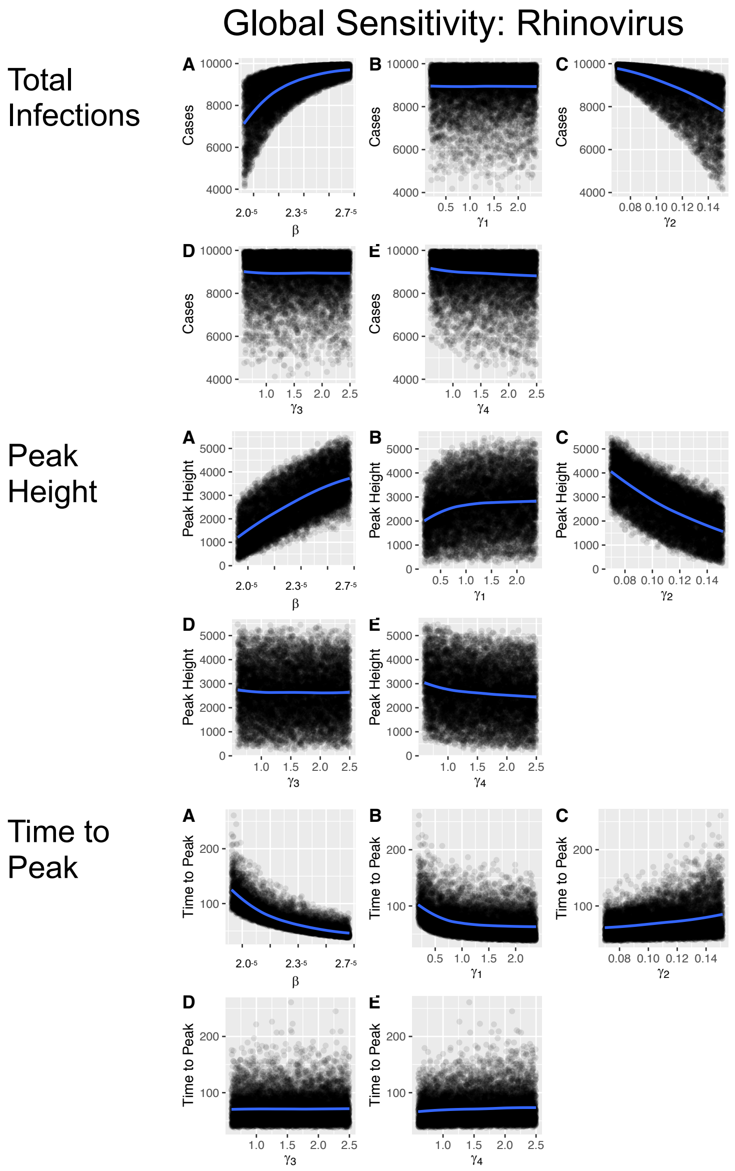


Figure E.9: Global sensitivity analysis for rhinovirus. Sub-plots A, B, C, D, and E show the distribution of 10,000 Latin Hypercube Samples for input variables $\beta, \gamma_1, \gamma_2, \gamma_3$, and $\gamma_4$ respectively. Local Polynomial Regression trend lines show their relationship to response variables total infections at the top, peak height in the middle, and time to peak at the bottom.

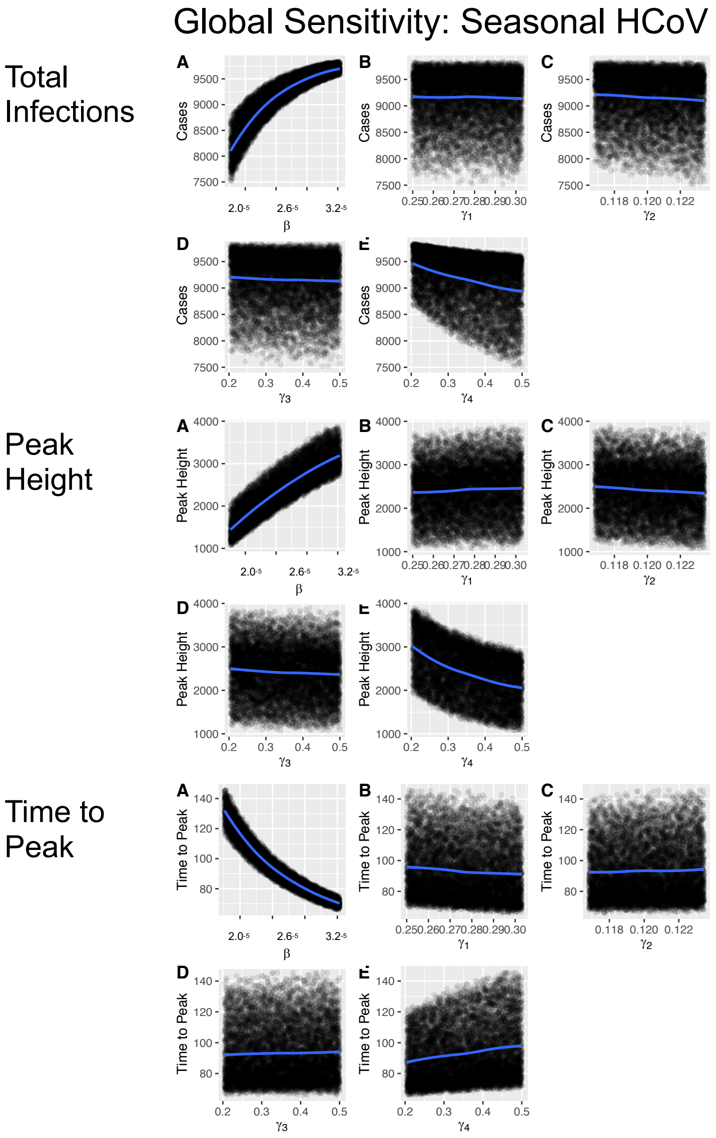


Figure E.10: Global sensitivity analysis for seasonal human coronavirus. Sub-plots A, B, C, D, and E show the distribution of 10,000 Latin Hypercube Samples for input variables $\beta, \gamma_1, \gamma_2, \gamma_3$, and $\gamma_4$ respectively. Local Polynomial Regression trend lines show their relationship to response variables total infections at the top, peak height in the middle, and time to peak at the bottom.

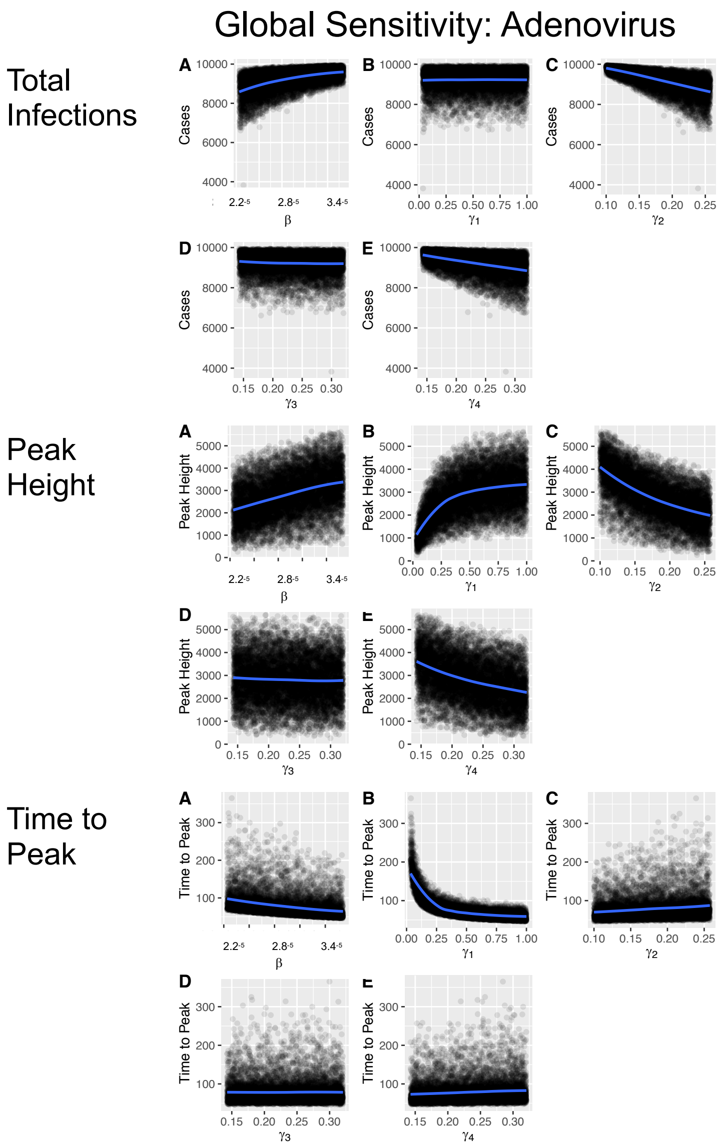


Figure E.11: Global sensitivity analysis for adenovirus. Sub-plots A, B, C, D, and E show the distribution of 10,000 Latin Hypercube Samples for input variables $\beta, \gamma_1, \gamma_2, \gamma_3$, and $\gamma_4$ respectively. Local Polynomial Regression trend lines show their relationship to response variables total infections at the top, peak height in the middle, and time to peak at the bottom.

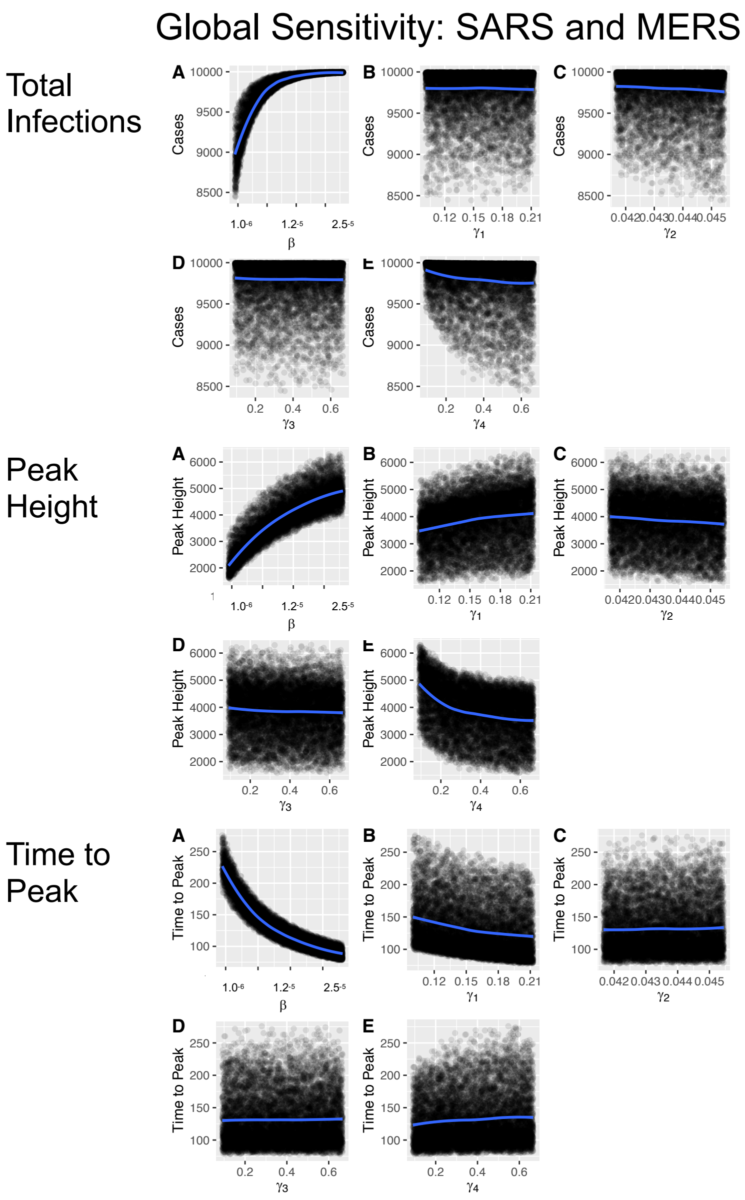


Figure E.12: Global sensitivity analysis for the historic coronavirus outbreak viruses, SARS and MERS. Sub-plots A, B, C, D, and E show the distribution of 10,000 Latin Hypercube Samples for input variables $\beta, \gamma_1, \gamma_2, \gamma_3$, and $\gamma_4$ respectively. Local Polynomial Regression trend lines show their relationship to response variables total infections at the top, peak height in the middle, and time to peak at the bottom.
